# Country differences in transmissibility, age distribution and case-fatality of SARS-CoV-2: a global ecological analysis

**DOI:** 10.1101/2021.02.17.21251839

**Authors:** Caroline Favas, Prudence Jarrett, Ruwan Ratnayake, Oliver J Watson, Francesco Checchi

## Abstract

**Introduction:** SARS-CoV-2 has spread rapidly across the world yet the first pandemic waves in many low-income countries appeared milder than initially forecasted through mathematical models. Hypotheses for this observed difference include under-ascertainment of cases and deaths, country population age structure, and immune modulation secondary to exposure to endemic parasitic infections. We conducted a country-level ecological study to describe patterns in key SARS-CoV-2 outcomes by country and region and to explore possible associations of the potential explanatory factors with these outcomes.

**Methods:** We collected publicly available data at country level and compared them using standardisation techniques. We then explored the association between exposures and outcomes using alternative approaches: random forest (RF) regression and linear (LM) regression. We adjusted for potential confounders and plausible effect modifications.

**Results:** Altogether, data on the mean time-varying reproduction number (mean *R_t_*) were available for 153 countries, but standardised averages for the age of cases and deaths and for the case-fatality ratio (CFR) could only be computed for 61, 39 and 31 countries respectively. While mean *R_t_* was highest in the WHO Europe and Americas regions, median age of death was lower in the Africa region even after standardisation, with broadly similar CFR. Population age was strongly associated with mean *R_t_* and the age-standardised median age of observed cases and deaths in both RF and LM models. The models highlighted other plausible roles of population density, testing intensity and co-morbidity prevalence, but yielded uncertain results as regards exposure to common parasitic infections.

**Conclusions:** The average age of a population seems to be an important country-level factor explaining both transmissibility and the median age of observed cases and deaths, even after age-standardisation. Potential associations between endemic infections and COVID-19 are worthy of further exploration but seem unlikely, from this analysis, to be key drivers of the variation in observed COVID-19 epidemic trends. Our study was limited by the availability of outcome data and its causally uncertain ecological design, with the observed distribution of age amongst reported cases and deaths suggesting key differences in surveillance and testing strategy and capacity by country and the representativeness of case reporting of infection. Research at subnational and individual level is needed to explore hypotheses further.

## Introduction

Since the end of 2019, the Coronavirus Disease 2019 (COVID-19), caused by the novel coronavirus SARS-CoV-2, has spread rapidly across the world, resulting in considerable morbidity and mortality, with more than 107 million cases and 2.37 million deaths reported globally as of February 11, 2021 (1).

Nevertheless, the COVID-19 pandemic has affected countries differently, with marked geographical disparities in the observed burden of cases and associated deaths. While the American continent bears the highest burden of cases and fatalities ascertained to date, African countries seem relatively spared, making up 2.5% of global cases and 2.9% of the global death toll despite accounting for 14% of the global population (2, 3). Indeed, the first pandemic waves in many low-income countries (LICs) appeared milder than initially forecasted (4–8).

Many hypotheses have been put forward to explain country differences in epidemic trajectory and magnitude. Weaker health systems with inequities in access and limited testing capacity may have resulted in the under-ascertainment of cases and deaths, the degree of which is uncertain (9).The forewarning from other health systems that were quickly overwhelmed in China and Europe may have led to comparatively earlier introduction of control measures relative to SARS-C-V-2 introduction in some LICs, increased stringency in application of these measures, and thus partial suppression of community transmission (10, 11).

In LICs, a younger population age structure could have had implications for virus transmission as well as having lower the infection fatality ratios due to a smaller proportion of older individuals who are most vulnerable to severe disease (12–14). In addition, efforts to shield older individuals in settings with younger populations may have been more effective due to a lower absolute number of individuals needing to be shielded. However, the increasingly multi-generational and larger households observed in less wealthy countries may offset this effect and increase the risk of exposure to older individuals (15). Furthermore, populations may have a lower prevalence of comorbidities that increase the risk of death with COVID-19 (16).

It has also been postulated that greater lifetime exposure to common infections in LIC populations may confer some immune protection from SARS-CoV-2 through a more diverse and competitive microbiome, more effective non-specific immune response and decreased likelihood of the ‘cytokine storm’ seen in severe COVID-19 disease (10, 17). Parasites such as *Plasmodium spp.* and soil-transmitted helminths, to which many LIC populations are exposed since early childhood, have immunomodulatory effects (18, 19). Access to improved water and sanitation may be a distal factor related to exposure to these parasites. Population density and household size are typical drivers of person-to-person disease transmission (20, 21).

There is little evidence for the relative influence of these hypothesised factors on the observed heterogeneity in epidemic trends around the world. In Figure 1, we propose a causal framework for understanding the relationship between potential explanatory factors, including those related to the hypotheses described above, and the outcomes of transmissibility, age-distribution of cases and deaths, and case-fatality of SARS-CoV-2 at the population level.

**Figure 1:**
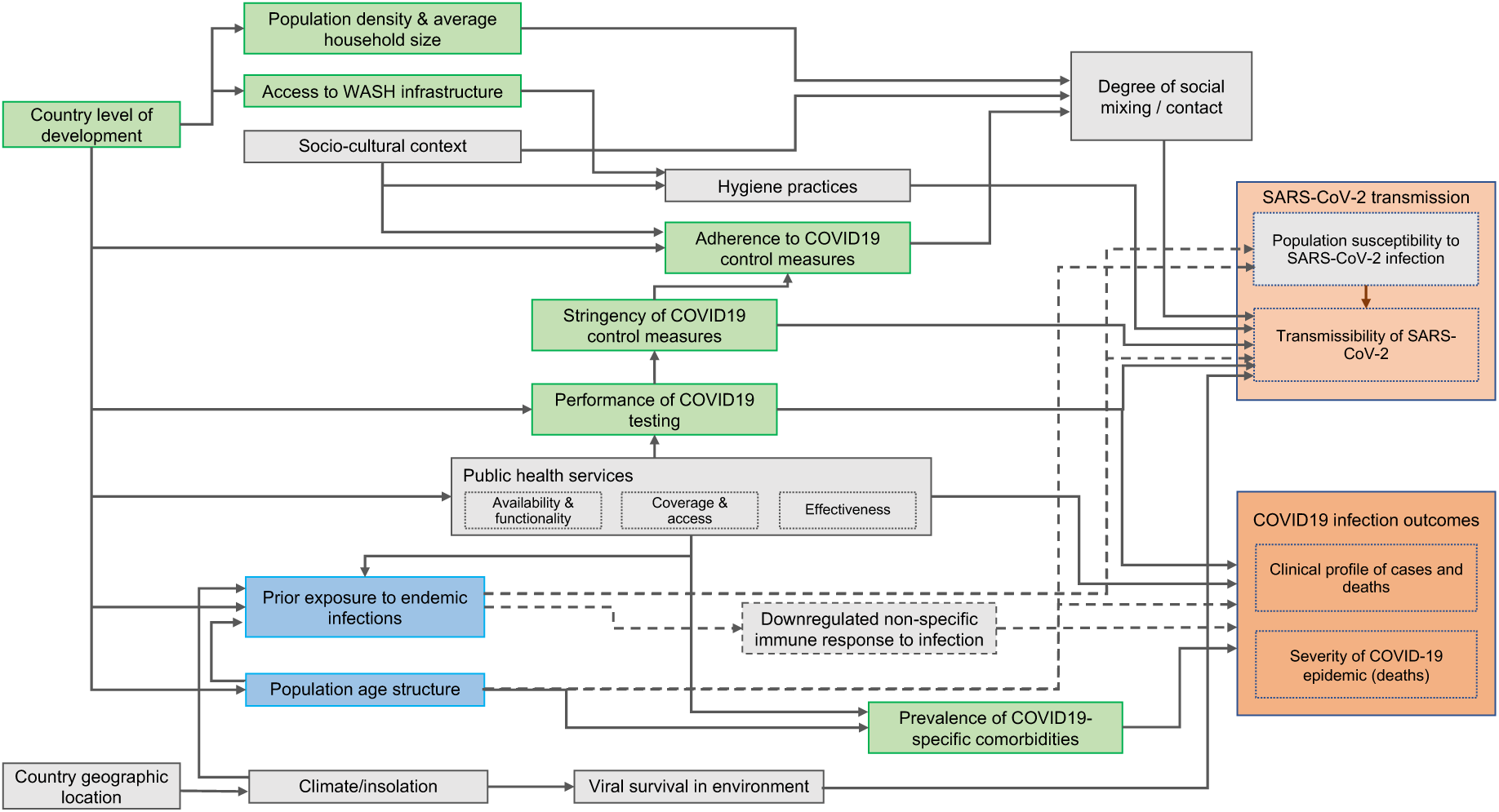
Proposed causal framework of factors determining SARS-CoV-2 transmissibility and COVID-19 disease outcomes. Boxes coloured in pink=outcome variables; boxes coloured in blue=exposures of interest; boxes coloured in green=covariates for which we obtained data; boxes coloured in grey=covariates and intermediate outcome variables for which we didn’t obtain data. Dotted lines represent hypotheses explored in this study.

We conducted a country-level ecological study to describe patterns in key SARS-CoV-2 outcomes by country and region, and explore possible associations of these outcomes with potential explanatory factors.

## Methods

### Study population and period

We considered data from each country in the world, as available; observed COVID-19 burden data from March to October 2020 were included. Analysis units included 193 United Nations member states plus the State of Palestine, Holy See and Hong Kong Special Administrative Region. Dependent territories and other entities were excluded due to inconsistencies in reporting.

### Independent variables

We sought publicly available data on indicators plausibly representing each of the domains in the causal framework (e.g., the Human Development Index was used for the level of development of a country; domains for which we could not identify suitable indicators are coloured in grey in Figure 1). Information on the indicators used and data sources is summarised in Table 1 and detailed in the Supplementary File.

**Table 1.** Summary of included variables, indicators and sources of data.

| Type of variable | Variable measured | Indicator | Year | Countries included | Data source |
| --- | --- | --- | --- | --- | --- |
| Outcome | Transmissibility of SARS-Cov-2 | Average reproduction number ($R_t$) estimates over the study time period, from the day when 50 cumulative deaths were reported | 2020 | 153 | Imperial College COVID-19 LMIC Reports (22) |
|  | Clinical profile of cases | Standardised median age of cases | 2020 | 61 | NA* |
|  | Clinical profile of deaths | Standardised median age of deaths | 2020 | 39 | NA* |
|  | Severity of COVID-19 epidemic | Observed case fatality ratio (CFR):<br>- Crude CFR<br>- Age-standardised CFR<br>- Incidence standardised CFR | 2020 | 169<br>31<br>31 | NA* |
| Independent | Prior exposure to infections: malaria | The age standardised mean predicted parasite prevalence rate for *Plasmodium falciparum* (*Pf*) malaria for children 2-10 years old | 2017 | 176 | The Malaria Atlas Project database (25) |
|  | Prior exposure to infections: malaria | The age standardised mean predicted all-age parasite prevalence rate for *Plasmodium vivax* (*Pv*) malaria | 2017 | 163 | The Malaria Atlas Project database (26) |
|  | Prior exposure to infections: other parasites | All-age point prevalence of infection with:<br>- soil-transmitted helminths<br>- schistosomiasis<br>- lymphatic filariasis | 2017 | 186 | Global Burden of Disease Study (27) |
|  | Country age structure | Median age (in years) of the population | 2020 | 185 | World Population Prospect (23) |
|  | Country level of development | Human development index | 2018 | 188 | UN Development Programme database (28) |
|  | Population density | Population density, as the number of persons per square kilometre | 2020 | 196 | World Population Prospect (23) |
|  | Household size | The average number of usual residents (household members) per household | Variable | 149 | UN Database on Household Size and Composition (29) |

Table 1. Continued.
| Type of variable | Variable measured | Indicator | Year | Countries included | Data source |
| --- | --- | --- | --- | --- | --- |
| Independent | Access to WASH infrastructures | The proportion of people using safely managed sanitation services, as a percentage of population | 2017 | 88 | World Bank World Development Indicator database (30) |
|  | Stringency of COVID-19 control measures | Average score for stringency index from 01/01/2020 to 09/09/2020 | 2020 | 169 | Oxford COVID-19 Government Response Tracker (31) |
|  | Performance of COVID-19 testing | Average score for testing policy indicator from 01/01/2020 to 09/09/2020 | 2020 | 169 | Oxford COVID-19 Government Response Tracker (31) |
|  | Performance of COVID-19 testing | Testing rate over the study time period | 2020 | 120 | NA* |
|  | Adherence to COVID-19 control measures (change in mobility) | The percentage net change in mobility across four categories (1- Retail & Recreation, 2- Grocery & Pharmacy, 3- Transit stations, 4- Workplaces). Average calculated over the period from 15/02/2020 to 09/10/2020 | 2020 | 130 | Google Community Mobility Reports (32) |
|  | Prevalence of comorbidities | Age-standardised percentage of country populations at increased risk of severe COVID-19, defined as those with at least one underlying condition listed as “at increased risk” in guidelines from WHO and public health agencies in the United Kingdom and United States | 2020 | 183 | Clark et al. (16) |
| *\*Data not from a single source. Description of how these data were obtained is found in the Supplementary File.*<br>SARS-CoV-2= severe acute respiratory syndrome coronavirus 2; COVID-19=coronavirus disease 2019; WASH=water, sanitation and hygiene; NA= non applicable; UN=United Nations |  |  |  |  |  |

### Outcome variables

#### Transmissibility

We sourced time-varying reproduction numbers (R*_t_*) by country, as estimated on a real-time basis by the Imperial College London COVID-19 Response Team (22). A characteristic of these estimates is that they are informed by the dynamics of observed COVID-19 deaths rather than cases, which are less likely to be detected and more susceptible to fluctuations in ascertainment over time due to changes in testing regimens. We averaged R*_t_* estimates over our analysis period (March-October), from the day when 50 cumulative deaths were reported. This was to ensure that these averages were not overly influenced by the prior distribution for R*_t_* used to inform the Bayesian framework for R*_t_* estimation (itself highly dependent on observations during the first days and weeks of observed transmission).

#### Age of observed cases and deaths

We conducted a systematic search of each country’s national COVID-19 website (e.g., Ministry of Health dashboard) or regional surveillance reports (e.g., produced by the African Centres for Disease Control and Prevention) for overall and age-stratified COVID-19 case and death data. Age-specific data on cases were available from 61 countries and age-specific data on deaths from 39 countries; 35 countries reported both values. For each country we present a “standardised median age” indicator interpretable as the median age of cases or deaths if the country’s observed age-specific cumulative incidence or death rates were applied to the world’s population age structure (23). Further detail can be found in Supplementary File 1.

#### Case-fatality ratio

After omitting countries with <50 total observed cases, we computed a crude case-fatality ratio (CFR) for each country by dividing observed deaths by cases. For countries with available age-specific data, we computed (i) an ‘age-standardised’ CFR, derived as above by applying countries’ age-specific crude CFRs to the world’s population structure; and (ii) an ‘incidence-standardised’ CFR, derived by applying each country’s age-specific CFRs to the observed age-specific caseload in South Korea, selected as a reference due to this country’s reportedly high coverage of case detection (i.e. relatively low selection bias affecting the profile of observed cases) and standard of care (24). The chosen standardisation method aims to (i) account for age differences in infection-fatality ratios (IFR), while (ii) reducing bias due to incomplete testing; neither, however, accounts for the effect of age structure on incidence or fully removes confounding.

### Statistical analyses

We present two approaches for exploring the associations of hypothesised exposures with each of the above outcomes, while adjusting for potential confounders and accounting for plausible effect modifications chosen a priori for each outcome. For mean R*_t_* and the crude CFR, we did a global analysis as well one specific to African countries (the data were too sparse for other outcomes to perform region-specific analyses). All analysis was done on R software (33). Data and R scripts are available at https://github.com/francescochecchi/covid_eco_analysis/.

#### Random forest regression

Random forest (RF) regression is a machine-learning approach that may be used to efficiently explore the importance of predictor variables, and possible effect modifications, for a given outcome. RF imposes minimal statistical assumptions on data, and copes well with collinearity (34). It consists of generating a large number of regression trees (i.e. partitions of the independent variables, in varying order, with each variable generating a node or ‘split’) and averaging over these based on their accuracy in predicting the outcomes. We implemented two RF approaches for each outcome, using the `randomForest` R package, with 1,000 trees grown: (i) using non-missing independent variable data only; (ii) imputing missing independent variable data through the `rfImpute` proximity method (35) (only variables with at least 60% completeness were subjected to imputation; remaining variables were excluded altogether). As the two approaches yielded similar results, for brevity we only present the latter. We then computed various metrics of variable importance, among which we present and consider most informative the mean minimal depth (MMD: a low value indicates that the variable is generally close to the root of the grown trees, i.e. a large proportion of the data are meaningfully partitioned on the basis of this variable); the mean squared error (MSE) increase (i.e. by how much model error increases if the variable is omitted); and the number of trees (out of 1000) in which the variable is the first node based on which the data are split (the higher, the more fundamental the variable may be).

#### Linear regression

As each outcome was continuous and not structured hierarchically, we applied ordinary least-squares fixed-effects linear models (LM), guided by our *a priori* causal framework, to explore associations. We imputed missing data for the independent variables using the `mice` package; as with the RF models, only variables with at least 60% completeness were subjected to imputation; variables were otherwise excluded. For each outcome, we first observed collinearity among independent variables through scatterplots and Pearson correlation coefficients (see Supplementary File). We screened potential confounding variables through univariate analysis (retaining variables with a *p* value < 0.20). We fit models through stepwise backward variable selection, retaining variables that improved goodness of fit (adjusted *R^2^* and F-statistic p-value testing whether the model fits data better than the null model) or influenced the effect of potential exposures on the outcomes. We tried alternative collinear variables and tested plausible two-way interactions. We verified model assumptions including normality and the homoscedasticity of residuals. For each outcome, we present two models: one with all exposures retained (Supplementary File); and a “reduced” model with only significant (*p* < 0.05) and/or model-influential exposures retained (Table 2).

**Table 2.** Summary of key associations between independent variables and the outcomes. Exposures of interest are in italics.

| Most important variables | Random forest regression |  |  | Multivariate linear regression (reduced model) |  |  |
| --- | --- | --- | --- | --- | --- | --- |
|  | MMD | MSE increase | Times a root | Coef. | 95% CI | p-value |
| **Outcome: mean time-varying reproductive number (Rt)** |  |  |  |  |  |  |
| *Median population age (years)* | 2.824 | 0.0056 | 62 | 0.0080 | 0.0049 to 0.0112 | <0.0001 |
| *Prevalence of lymphatic filariasis (%)* | 2.564 | 0.0046 | 131 | -1.9168 | -3.1422 to -0.6915 | 0.0024 |
| *Prevalence of Pf (%)* | 3.122 | 0.0029 | 66 | - | - | - |
| Mean household size (persons) | 2.202 | 0.0093 | 118 | - | - | - |
| Mean mobility change (%) | 2.358 | 0.0025 | 32 | - | - | - |
| Population density (persons per square kilometre) | 2.996 | 0.0015 | 2 | -0.000036 | -0.000060 to -0.000012 | 0.0041 |
| Mean stringency index (score) | 3.472 | 0.0013 | 0 | 0.0023 | 0.0003 to 0.0043 | 0.0228 |
| **Main effect modifications** | **MMD** | **Occurrences** |  | **Coef. %** | **95% CI** | **p-value** |
| Median population age x testing policy | 2.195 | 323 |  | - | - | - |
| Median population age x testing rate | 3.130 | 226 |  | - | - | - |
| (Adjusted)<sup>1</sup> R-squared (F-test; p-value) |  | 0.38 |  |  | 0.31 (F = 16.88; p < 0.0001) |  |
| **Outcome: age-standardised median age of observed cases** |  |  |  |  |  |  |
| *Median population age (years)* | 2.144 | 6.2533 | 106 | -0.3961 | -0.5276 to -0.2645 | <0.0001 |
| *Prevalence of STH (%)* | 3.179 | 2.2431 | 80 | - | - | - |
| Mean mobility change (%) | 2.537 | 1.4986 | 28 | - | - | - |
| -29 to -20 | - | - | - | 1.6983 | -0.9826 to 4.3793 | 0.2097 |
| -19 to -10 | - | - | - | 0.7165 | -1.9888 to 3.4218 | 0.5978 |
| -10 or less | - | - | - | 4.0711 | 0.8766 to 7.2657 | 0.0134 |
| Mean testing rate per population (per 1,000) | 2.434 | 2.0958 | 76 | - | - | - |
| Population at increased risk (%) | 2.169 | 4.0191 | 85 | - | - | - |
| **Main effect modifications** | **MMD** | **Occurrences** |  | **Coef. %** | **95% CI** | **p-value** |
| Median population age x mean mobility change | 2.210 | 206 |  | - | - | - |
| Median population age x mean stringency index | 1.980 | 201 |  | - | - | - |
| Median population age x proportion at increased risk | 1.376 | 251 |  | - | - | - |
| (Adjusted)<sup>1</sup> R-squared (F-test; p-value) |  | 0.25 |  |  | 0.44 (F = 12.58; p < 0.0001) |  |
| **Outcome: age-standardised median age of observed deaths** |  |  |  |  |  |  |
| *Median population age (years)* | 1.768 | 9.8945 | 119 | 0.3974 | 0.1702 to 0.6245 | 0.0011 |
| *Prevalence of STH (%)* | 3.208 | 5.5241 | 87 | - | - | - |
| Mean stringency index (score) | 1.848 | 8.9399 | 92 | -0.2044 | -0.3579 to -0.0509 | 0.0105 |
| Mean testing rate per population (per 1,000) | 1.946 | 6.0048 | 71 | - | - | - |
| Population at increased risk (%) | 1.727 | 6.5484 | 68 | -0.4517 | -0.8482 to -0.0553 | 0.0267 |
| **Main effect modifications** | **MMD** | **Occurrences** |  | **Coef. %** | **95% CI** | **P-value** |
| Median population age x mean mobility change | 2.005 | 105 |  | - | - | - |
| Median population age x mean stringency index | 1.496 | 171 |  | - | - | - |
| Median population age x proportion at increased risk | 1.923 | 162 |  | - | - | - |
| (Adjusted)<sup>1</sup> R-squared (F-test; p-value) |  | 0.63 |  |  | 0.65 (F = 24.53; p < 0.0001) |  |
| **Outcome: age-standardised CFR** |  |  |  |  |  |  |
| *Median population age (years)* | 1.851 | 0.0000 | 105 | - | - | - |
| Population at increased risk (%) | 2.403 | 0.0000 | 68 | 0.0814 | 0.0159 to 0.1470 | 0.0167 |
| **Main effect modifications** | **MMD** | **Occurrences** |  | **Coef. %** | **95% CI** | **P-value** |
| Median population age x mean stringency index | 0.930 | 128 |  | - | - | - |
| Median population age x proportion at increased risk | - | - |  | -0.0019 | -0.0041 to 0.0002 | 0.0787 |
| (Adjusted)<sup>1</sup> R-squared (F-test; p-value) |  | -0.21 |  |  | 0.19 (F = 4.63 p < 0.05) |  |

<sup>1</sup> for LM only.
Abbreviations: CFR= case fatality ratio; CI= confidence interval; Coef.= coefficient; MMD= mean minimal depth; Pf= Plasmodium falciparum; Pv= Plasmodium Vivax; STH= soil-transmitted helminths; yo= years old

## Results

### Observed country patterns

Completeness for mean *R_t_* and CFR was >70% for all regions except WPRO (33.3% and 51.9%, respectively); completeness for median age of cases and deaths was much lower, ranging from 13.6% (EMRO) to 52% (EURO), and 0 (SEARO) to 36% (EURO), respectively (Supplementary File). Figure 2 summarises trends in each of the outcomes by WHO region, as available. Mean *R_t_* was highest in the EURO and PAHO regions (range 0.92 to 1.77 and 0.73 to 1.73, respectively) and lowest in the AFRO region (0.96 to 1.45). The age distribution of COVID-19 cases and deaths was heterogeneous across regions: even when standardised for differences in age structure, the median age of observed cases was higher in the AFRO and EMRO regions, whereas ascertained deaths occurred at younger ages in those regions compared to EURO and WPRO. While the crude CFR did not seem to vary widely across regions, higher CFR was found in the EMRO, AFRO, and PAHO regions when CFR was standardised for age and incidence. Figure 3 compares the age-standardised median age of cases and deaths for each country. Countries in the EURO and PAHO regions are clustered in the upper left quadrant (i.e., median age of cases <40, median age of deaths >70). Most countries in the AFRO region are clustered in the lower right quadrants (i.e., median age of cases >40, median age of death <70). The Supplementary File provides results by country and graphical explorations of the correlation between independent variables and outcomes.

**Figure 2.**
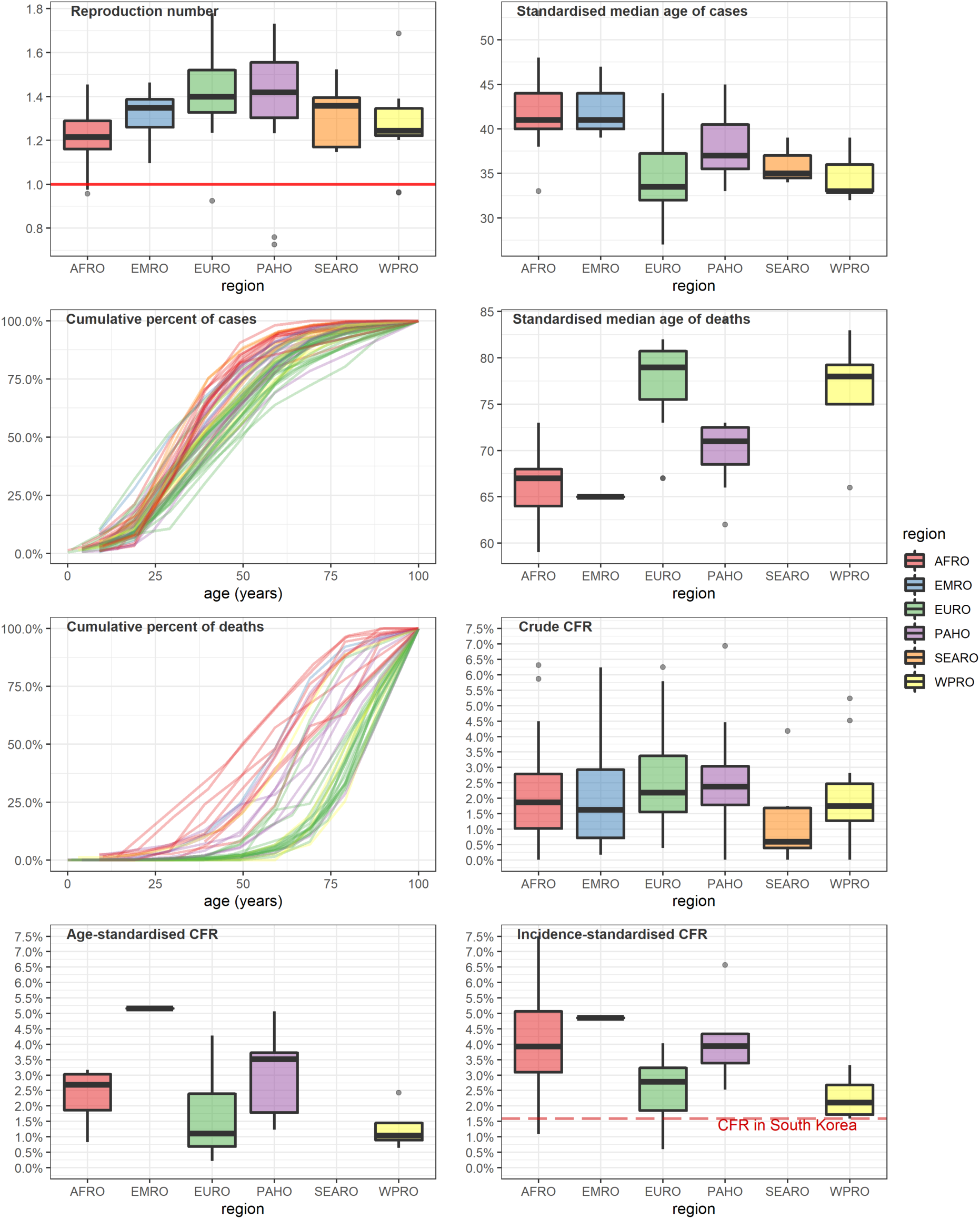
Analysis outcomes, by WHO region. All boxplots indicate the median and inter-quartile range (boxes), 95% percentile intervals (whiskers) and outliers (dots). CFR = case-fatality ratio. AFRO= African regional office; EMRO= Eastern Mediterranean regional office; EURO= European regional office; PAHO= Pan American health organisation; SEARO= South-East Asia regional office; WPRO= Western Pacific regional office

**Figure 3.**
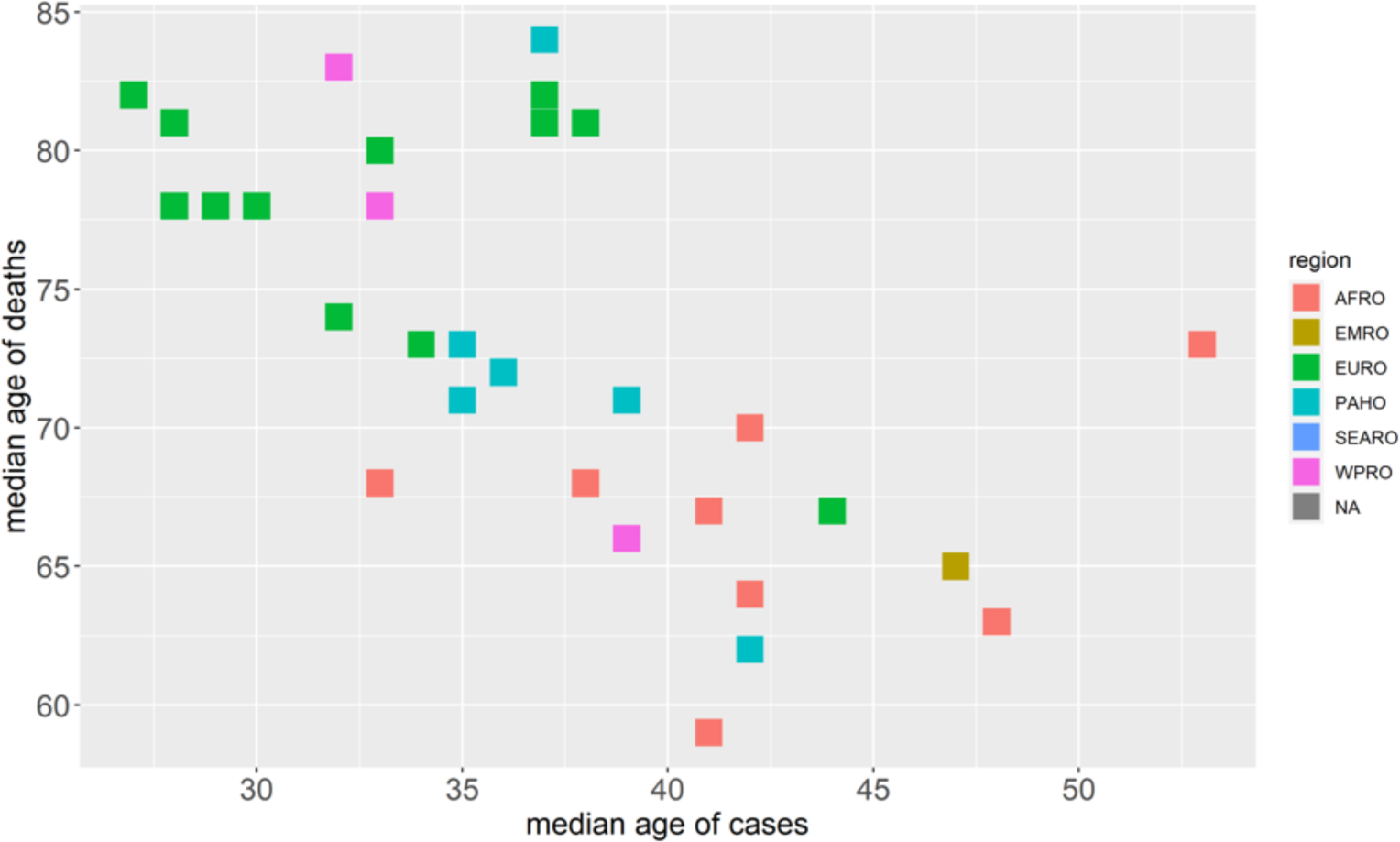
Scatter plot diagram of the age-standardised median age of deaths and cases (in years) for 35 countries for which both could be computed.

### Statistical associations

Table 2 summarises key results from the two multivariate regression models (RF and the reduced versions of the LMs) of imputed predictors for mean R*_t_,* age-standardised median age of observed cases, age-standardised median age of observed deaths, and age-standardised CFR. Models for crude and incidence-standardised CFR were excluded as their fit was poor. The Supplementary File presents detailed results (all exposures fitted) for each outcome globally, the AFRO region for transmissibility and crude CFR, and the models where mean *R_t_* is computed after ≥50 cumulative deaths.

In the RF model for mean *R_t_*, mean household size, prevalence of filariasis and median population age were the three most important variables when considering the different metrics of importance. Mean mobility change, population density, and prevalence of *Pf* were also important. Testing rate and testing policy were effect modifiers for the association between median population age and mean *R_t_*. In the LM model, prevalence of filariasis, median population age, mean stringency and population density showed significant associations (p<0.05). The association with *Pf* prevalence was non-significant in the reduced model. When considering only the AFRO region, population age, population density, and mean stringency did not remain important, but the importance of prevalence of filariasis increased in the RF model and remained significantly associated in the reduced LM model (p<0.001).

In the RF models for median age of observed cases and deaths, median population age was the most important variable (i.e. even after age-standardisation), along with proportion at increased COVID-19 risk, testing rate, and prevalence of helminths. Mobility change was also important for median age of observed cases whereas stringency index was important for median age of observed deaths. Mean mobility change, mean stringency index, and proportion at increased risk were effect modifiers. In the LM model, median population age was positively associated with median age of cases (p<0.0001) and negatively associated with median age of deaths (p<0.01). The prevalence of helminths was not significantly associated with either outcome.

Lastly, RF suggested that median population age was also an important predictor of age-standardised CFR, but this was not borne out in the LM. Both models had a poor fit.

## Discussion

We aimed to identify important factors at national level which may explain the global heterogeneity of SARS-CoV-2 epidemics and, specifically, to explore the role of population age structure and exposure to endemic infections. We found that median population age is an important factor explaining variability in transmissibility and the age of observed cases and deaths at the national level, with a significant association remaining even after age-standardisation. Potential associations between endemic infections and COVID-19 appear unlikely, based on this analysis alone, to be key drivers in the variation in observed COVID-19 epidemic trends, though the association with filariasis prevalence at global and AFRO level is intriguing (below). Furthermore, the observed distribution of age amongst reported cases and deaths (after age-standardisation) at least suggests key differences in surveillance and testing capacity between countries and regions, which probably affect the representativeness of reported cases and deaths. Overall, while we emphasise caution over causal inference due to possible hidden confounding and incomplete data, we find that population age structure presents a consistent association that suggest its full impact on country-specific epidemics warrants further research.

### Population age structure

Similar to what has been observed with severe acute respiratory syndrome (SARS) and Middle East Respiratory Syndrome (MERS) coronaviruses (36), most studies of COVID-19 suggest that children are both less susceptible to infection and less infectious, as indicated by lower secondary attack rates from younger index cases (37–39). Our study provides some evidence that this may also play out at the population level such that countries with younger population age structure have a smaller susceptible population, less transmission and milder epidemics that may reflect epidemic trends in Sub-Saharan Africa in particular. However, epidemiological studies on the role of children have often relied on passive case detection and thus are likely to miss the majority of pauci-or asymptomatic cases in these age groups (40). Outbreaks among children and adolescents have been important in introducing transmission into households in the United Kingdom (41). Although refuted by some (13, 42), the role of secondary school-aged children (age 11-18 years) is considered an important driver of transmission by others (40). Heterogeneity in social contact patterns across age and locations may also influence the role population age structure plays in the transmission of SARS-CoV-2 and explain this discrepancy. Across Europe contacts between children and young people are highly assortative – that is, they mix most with people of a similar age-indicating their importance in the transmission of disease between households in these countries (43). Through modelling, Prem, Cook and Jit find that intergenerational contact was particularly common in South Asia, and that household contact patterns in countries with much younger population structures did not conform to those in Europe, being characterised by all age groups including adults having most contact with children and teenagers (44), such that the likelihood of a vulnerable older adult coming into contact with an infectious case (and thus the risk of onward transmission or death) may be smaller. Interestingly, in Kenya, assortative mixing was most pronounced between children in rural areas whereas in semi-urban areas it was most marked among adults (45). Therefore, country differences in urbanization and urban demography may reduce the importance of school-aged children in transmission, in line with our findings.

When restricting analysis to the AFRO region, population age structure did not remain an important variable for the prediction of transmission, and testing variables were less important, likely reflecting increased homogeneity in both age structure and lower testing capacity across African countries reducing our ability to detect significant associations.

Older population age structure was associated with lower median age of COVID-19 cases, after standardising for age and adjusting for confounding. Descriptively, the median ages of cases and deaths tended to be respectively ≥40 years and ≤70 years in the AFRO countries, as compared to countries in the EURO and PAHO regions. This contradicts what is known about age-dependent risk of symptomatic COVID-19. Control measures or behaviour change strategies targeting older people in countries with younger populations may explain this observation. Neither stringency index nor change in mobility are disaggregated by age. Alternatively, higher-income countries with older populations may have tested more young people due to greater testing capacity, while countries with low testing capacity might have prioritised older and at-risk people for testing, or reserved testing for travellers. Similar patterns were observed in higher income countries earlier in the pandemic when testing was not widely used and focussed on diagnosing severe infections and infections in key workers in order to assist with quarantine efforts (46). Testing rate and policy appear to modify the effect of age, supporting this explanation. However, age structure retained its importance even after including these effect modifications.

Conversely, countries with older populations had a higher median age of observed deaths from COVID-19, even after age-standardisation. Taking into consideration the fact that these same countries have younger cases overall, these findings may reflect better clinical care in countries with older populations so that although younger people are more likely to be infected, they are also much more likely to survive severe disease due to high-quality inpatient and intensive care. Moreover, outbreaks in nursing and long-term care settings have accounted for a large proportion of deaths in high-income countries and disproportionately affected older people (47). Notably, increasing prevalence of comorbidities was associated with younger age-standardised age of death, which may reflect a phenomenon whereby countries with a comparatively higher prevalence of diabetes, cardiovascular disease and other chronic conditions are also those where these conditions occur at a younger age due to life-course risk factors..

### Prior exposure to endemic infections

For transmissibility, prevalence of filariasis was the only measure of endemic infection that ranked highly in the RF model and that showed a strong negative association in the LM model. A plausible explanation may be that country prevalence of filariasis is actually a proxy measure for an unknown factor not included in our models. However, when considering the AFRO region only, where countries share more similar characteristics, the association between prevalence of filariasis and transmissibility becomes stronger. One possible, albeit tenuous biological mechanism relates to the fact that individuals with prior microfilarial infection appear to develop long-term or lifelong deficiency in IgA antibodies, which in turn may be associated with a lower proinflammatory response induced by Th1-type cytokines (49). In SARS-CoV-2 infections, the immunological response involving T-cells seems to be skewed towards these Th1 cells, especially in patients with severe disease (50). Therefore, prior exposure to filariasis may reduce the probability of individuals infected by SARS-CoV-2 to become symptomatic, which in turn may lower their infectiousness and thus population-level transmissibility.

Prevalence of *Pf* was also a variable of importance, albeit moderate, for the prediction of transmissibility in the RF model. However, the association was not significant in the reduced LM model. Although results from the RF model do not indicate a direction of association, one hypothesis that emerged from the literature was about cross reactivity: prior exposure to malaria triggers the production of poly-specific antibodies capable of interacting with multiple antigens, which may confer some protection against SARS-CoV-2 infection (47).

It is plausible that comparison at national level is insufficiently granular to detect any potential effect of endemic infections. Mbow et al (2014) report within-country variation in non-specific immunity when comparing urban and rural populations that could be related to changes in exposure to infections (51). Our analysis does not reflect this heterogeneity and sub-national data were not publicly available for the vast majority of countries. For soil-transmitted helminths in particular, contradictory hypotheses have been formulated. One of them stipulates that helminths could worsen COVID-19 severity, due to their immunomodulatory effects involving type 2 immune response (52, 53). An alternative hypothesis is that pre-exposure to helminths infection could reduce the risk of severe COVID-19 outcomes, through a downregulation in the production of proinflammatory cytokines involved in the hyperinflammation of the lungs and ‘cytokine storm’ commonly observed in severe and fatal COVID-19 disease (18, 19).

### Case-fatality

Our descriptive analysis does not indicate a lower CFR in the AFRO region compared to elsewhere, which does not support a narrative that the virus is less lethal in this region. Nonetheless, the low number of country observations for the age- and incidence-standardised CFR models is a limitation, and findings related to CFR are likely subject to significant confounding by poor case ascertainment. Generally, the CFR models fit poorly. The impact of a large number of undiagnosed cases on the CFR, and the limitations of its use in an ongoing epidemic are well-known (54). While it is logical that population age is an important determining factor, we cannot conclude this on the basis of our results.

### Limitations

Related to the ecological design of our study, one of the most important limitations is incomplete ascertainment of cases and deaths due to surveillance quality, testing capacity, and cause-of-death ascertainment, which is likely to be highly-variable among countries. We attempted to control for confounding at the aggregate level, with respect to testing. We controlled for testing performance by adjusting for the population testing rate and testing policy but acknowledge that these measures are themselves subject to bias. Low levels of testing in many low-income countries persist, and may mask the true scale of the epidemic. Indeed, studies point to substantial under-ascertainment of deaths in some countries (55, 56). Furthermore, testing data that are available may overrepresent older and sicker patients. Testing data disaggregated by age, sex, socioeconomic status and geographic location were not publicly available in most countries so that it is not possible to estimate the extent to which case-ascertainment reflects bias in these factors. Therefore, our findings should be interpreted with caution. We also note that countries with lower human development index and economic indicators generally have higher prevalence of endemic infections. As such, we could not fully adjust for all confounders.

Our study is based on a causal framework describing an evolving and incompletely understood pandemic, reflecting the extent of scientific understanding of the disease at the time. Our conceptual model may therefore not take into account all factors that influence SARS-CoV-2 epidemic trends and residual confounding due to these unknown factors may exist.

In general, ecological studies are useful to generate hypotheses, but do not provide a basis for causal inference. For example, stringency of control measures appears associated with higher *R_t_*: this may simply reflect reverse causality, whereby countries with high observed transmission would have maintained strict measures for longer. We averaged values for those variables that change over time, which may obscure the temporal relationships between them and means we cannot draw conclusions about variations in epidemic trends over time. A longitudinal study based on the same variables would be a useful next step.

Lastly, for many of the independent variables, our data is derived from modelled estimates (e.g. prevalence of soil-transmitted helminths) which are themselves based on limited national level data and therefore may not reflect the true measure of this variable. Unsystematic error in explanatory data would have biased coefficients towards the null, and thus masked potential associations.

## Conclusions and further work

Population age structure appears to be an important factor associated with the transmissibility of SARS-CoV-2 and the age-distribution of COVID-19 cases and deaths at the national level, even after such outcomes are age-standardised. Our findings do not conclusively support an effect of exposure to endemic parasitic infections on either transmissibility or age distribution of cases and deaths. Research at subnational or individual level should be conducted to investigate these hypotheses further. Where possible, analysis considering the sociodemographic characteristics of those tested will be useful in understanding the general role of lifelong exposures to infection in the observed patterns of disease. Further, study of social contact patterns in a broader range of countries, and the role of urbanization could provide useful insights. This work may be important not only for SARS-CoV-2 but could also inform preparedness and response to future pandemic threats.

## Data Availability

For our ecological study, we used publicly available data only.
Data and R scripts are available at https://github.com/francescochecchi/covid_eco_analysis/

## Acknowledgements

We are grateful to colleagues at the Imperial College of London for supporting our work and for providing useful feedback on the draft of this study report, in particular Charlie Whittaker. We also acknowledge the contribution of John Ackers and Helena Helmby, who provided helpful insights on parasitic infections and their immune modulatory effects.

## SUPPLEMENTARY FILE

### Details on variables

**Details on independent variables**

#### Exposure to endemic infections

We selected prevalence of malaria, soil-transmitted helminths, filariasis and schistosomiasis as indicators of prior exposure to endemic infections based on the literature (18,19,48,57) and on consultation with parasitology experts. Prior exposure to malaria was measured using age-standardised predicted parasite prevalence rate for children two to ten years of age for *Plasmodium falciparum*, and in all ages for *Plasmodium vivax*. Parasite rate is a commonly used index of malaria transmission intensity and as such, endemicity (58). The Malaria Atlas Project database provided country-specific data for the year 2017 (25, 26). Data werewere obtained for 177 and 164 countries for Plasmodium falciparum and Plasmodium Vivax parasite rates respectively.

Estimates of national point prevalence of schistosomiasis, lymphatic filariasis and soil-transmitted helminths (STH -grouping ascariasis, trichuriasis and hookworm disease) were available for 185 countries from the Global Burden of Disease Study 2017 (27).

#### Population age structure

We used the population’s median age in years to reflect countries’ age structure. Data for the year 2020 were taken from the 2019 revision of World Population Prospect (23).

#### Country level of development

We abstracted country-specific data on the Human Development index (HDI) from the United Nations Development Programme database, for 188 countries for the year 2018 (28). HDI is a summary measure of achievement in three basic dimensions of human development: a long and healthy life, knowledge and a decent standard of living (59).

#### Population density, household size and WASH infrastructure

Country-level data about population density (as the number of persons per square kilometre) for the year 2020 was taken from the 2019 revision of World Population Prospect for 196 countries (23). We obtained data about household size (average numbers of members) from the United Nations Database on Household Size and Composition 2019 for 149 countries (29). The proportion of people using safely managed sanitation services for the year 2017 (% of population), was used as a proxy to measure the access to water, sanitation, and hygiene services and infrastructures. Data were extracted from the World Bank World Development Indicator database for 88 countries (30).

#### Testing rate and testing policy

We captured data for two indicators – “testing policy’ and “stringency index” -from the Oxford COVID-19 Government Response Tracker which systematically records individual policy responses for more than 180 countries (31). Country-specific strategies regarding availability of polymerase chain reaction (PCR) testing for COVID-19 are translated into a daily score ranging from 0 (no testing policy) to 3 (open public testing even to asymptomatic people) (60). We abstracted data for 169 countries, covering the period from January 1 to September 9, 2020 and calculated an average score for each country over that time period.

Testing rate was derived from the total number of tests for each country collected as described below under “Outcomes” and calculated based on the total population for each country as reported by the World Bank in 2019 (30).

#### Stringency of COVID-19 control measures

The Stringency Index measures the strictness of control measures that primarily restrict people’s behaviour over the course of the pandemic (31). This is a composite daily measure which combines nine individual indicators (school closing, workplace closing, cancellation of public events, restrictions on gatherings, closing of public transports, stay at home requirements, restrictions on internal movements, international travel controls, and public information campaign) taking into consideration whether they are targeted or applied to the whole population (60). We extracted data for 169 countries, covering the period from January 1, 2020 up to September 9, 2020. To obtain a single value, we averaged the country-specific daily indices for the study time period.

#### Adherence to COVID-19 control measures

Mobility data were sourced from the Google Community Mobility Reports (32) and reflects adherence to COVID-19 control measures through the daily change in population movement relative to the pre-pandemic baseline. It is reported across six place categories: ‘residential’, ‘parks’, ‘grocery and pharmacy’, ‘transit stations’, ‘retail and recreation’, ‘workplaces’. The latter four categories are understood to be most reflective of the impact of control measures, and as such we averaged them into a single aggregate change. We abstracted data for 130 countries between February 15, 2020 and September 9, 2020.

#### Prevalence of comorbidities

To reflect the prevalence of comorbidities, we used the estimates made by Clark et.al of the percentage of the population at increased risk of severe COVID-19 in each country, defined as those with at least one underlying condition listed as “at increased risk” in guidelines from WHO and public health agencies in the United Kingdom and United States, not adjusted for age (61). Data were obtained for 183 countries.

### Details on outcome variables

We used the European Centre for Disease Prevention and Control (ECDC) list of national COVID-19 data sources (linking to country coronavirus dashboards, Ministry of Health webpage or social media accounts) to collect the following: cumulative number of coronavirus cases to date; cumulative number of coronavirus deaths to date; total number of coronavirus tests completed to date. The most recently updated figures were used. Age-disaggregated counts of cases and deaths were collected where available. Where age-disaggregation was given as a percentage of the total rather than a count, numbers of cases and deaths were calculated based on the most recently reported cumulative number of cases or deaths respectively.

The ECDC list was downloaded on 30th July 2020 (62). If the sources listed did not contain these data, or a source was not provided, a manual internet search for the country Ministry of Health website or coronavirus dashboard was attempted. If data were still not found, they were retrieved from the Our World In Data repository (63).

The count and proportion of missing outcome data by region is described in the table below.

**Table S1:** completeness of outcome data by WHO region

| Outcome | AFRO<br>(n, %)<br>N=49 | EMRO<br>(n, %)<br>N=22 | EURO<br>(n, %)<br>N=50 | PAHO<br>(n, %)<br>N=39 | SEARO<br>(n, %)<br>N=11 | WPRO<br>(n, %)<br>N=27 |
| --- | --- | --- | --- | --- | --- | --- |
| $R_t$ mean | 38 (77.6) | 22 (100) | 48 (96) | 28 (71.8) | 8 (72.7) | 9 (33.3) |
| crude_CFR | 46 (93.9) | 22 (100) | 48 (96) | 30 (76.9) | 9 (81.8) | 14 (51.9) |
| median_age_cases | 13 (26.5) | 3 (13.6) | 24 (52) | 28 (48) | 3 (27.3) | 7 (25.9) |
| median_age_deaths | 9 (18.4) | 1 (4.5) | 18 (36) | 7 (17.9) | 0 | 4 (14.8) |

#### Reproduction number

We sourced time-varying reproduction numbers (*R_t_*) by country, as estimated on a real-time basis by the Imperial College London MRC Centre for Global Infectious Disease Analysis (22). A characteristic of these estimates is that they are informed by the evolution of observed deaths rather than cases. Data were downloaded on 11th October 2020. We averaged *R_t_* estimates over our analysis period, from the day when 50 cumulative deaths were reported.

#### Age of observed cases and deaths

Age-specific data on cases were available from 61 countries and age-specific deaths from 39 (35 countries reported both). Eleven separate systems for age categorisation were used across countries, rendering direct comparisons difficult. Moreover, we wished to analyse differences in the average age of cases and deaths not merely explainable by countries’ demographic structure. Accordingly, for each country we present a “standardised median age” indicator *â*_*s*_, interpretable as the median age of cases or deaths if the country’s observed age-specific cumulative incidence or death rates were applied to the world’s population age structure. The quantity is computed as follows. Suppose a given country reports *x_i_* cumulative cases (or deaths) in each age category *a_i_* = {*a_1_, a_2_, a_3_ … a_max_*}, where *a_max_* is censored at 100 years. We can calculate age-specific attack rates 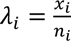, where, *n*_i_ is the population within each age category. If the country had the same population and age structure as the world, its expected age-specific caseload would be *w_i_ = λ_i_N_i_*, where *N*. denotes the world’s population. Accordingly, *â*_*s*_ is the age at which the 50th percentile of the cumulative sum of {*w_i_ … w_max_*} occurs. Since age increments in our dataset were not annual, we linearly interpolated *w_i_* over the range *a* = {0,100} before computing the median. We sourced projected age-specific population for each country and the world in 2020 from the United Nations World Population Prospects (23).

#### Case-fatality ratio

International comparisons of the lethality of SARS-CoV-2 are notoriously confounded by varying testing coverage, age distribution of cases, prevalence of co-morbidities and other factors. After omitting countries with < 50 total observed cases, we computed a crude case-fatality ratio (CFR) for each country by dividing observed deaths by cases. For countries with available age-specific data we also computed (i) an ‘age-standardised’ CFR, derived as above by applying countries’ age-specific crude CFRs to the world’s population structure; and (ii) an ‘incidence-standardised’ CFR derived by applying each country’s age-specific CFRs to the observed age-specific caseload in South Korea, selected as a reference due to this country’s reportedly high coverage of case detection (i.e. relatively low selection bias affecting the profile of observed cases) and standard of care. Neither standardisation method fully removes confounding; (i) accounts for age differences in infection-fatality ratios (IFR) and (ii) reduces bias due to incomplete testing, but neither accounts for the effect of age on incidence.

## Descriptive tables and graphs

**Figure S1.**
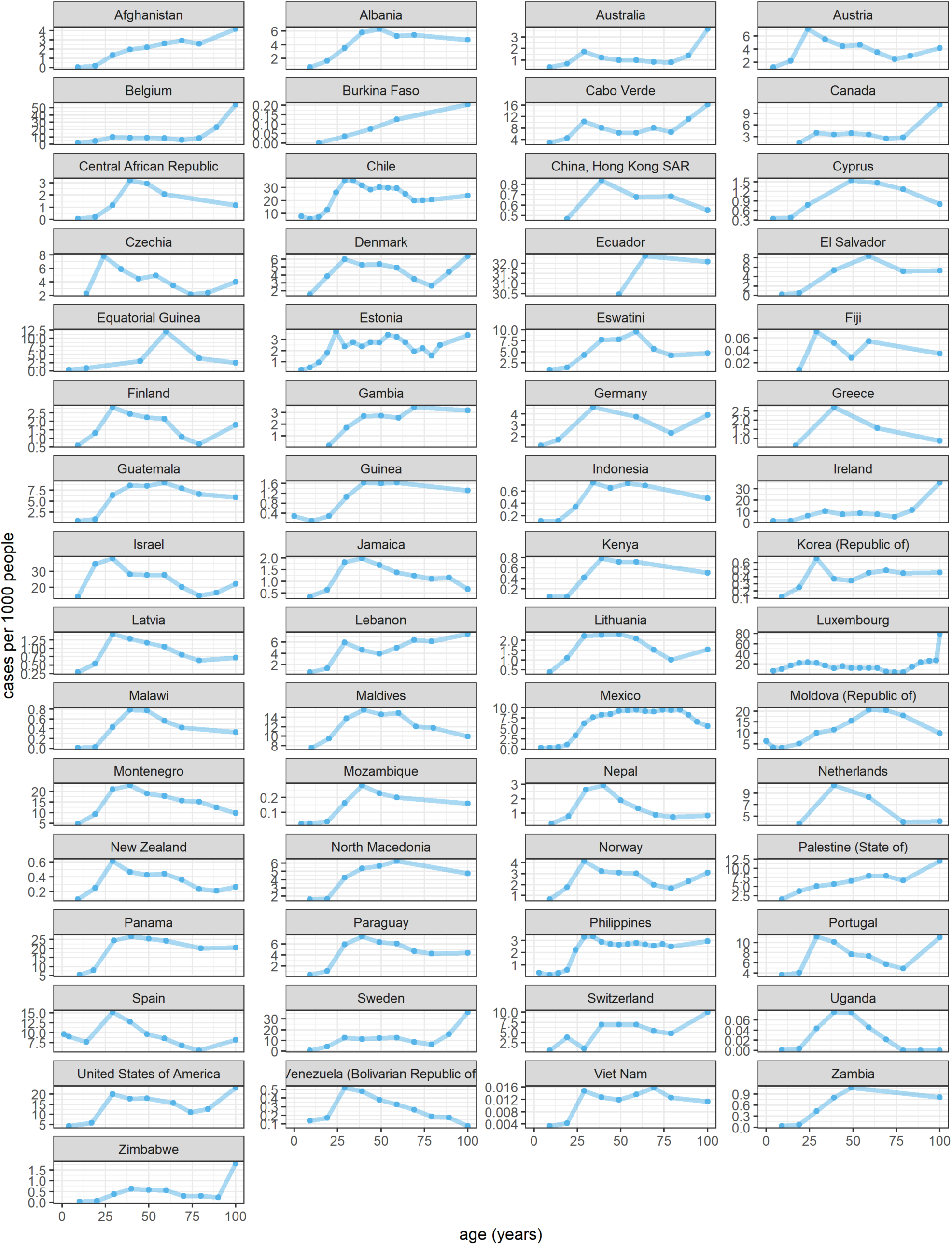
Age-specific cumulative incidence rate, by country or territory (observed COVID-19 cases per 1000 population).

**Figure S2.**
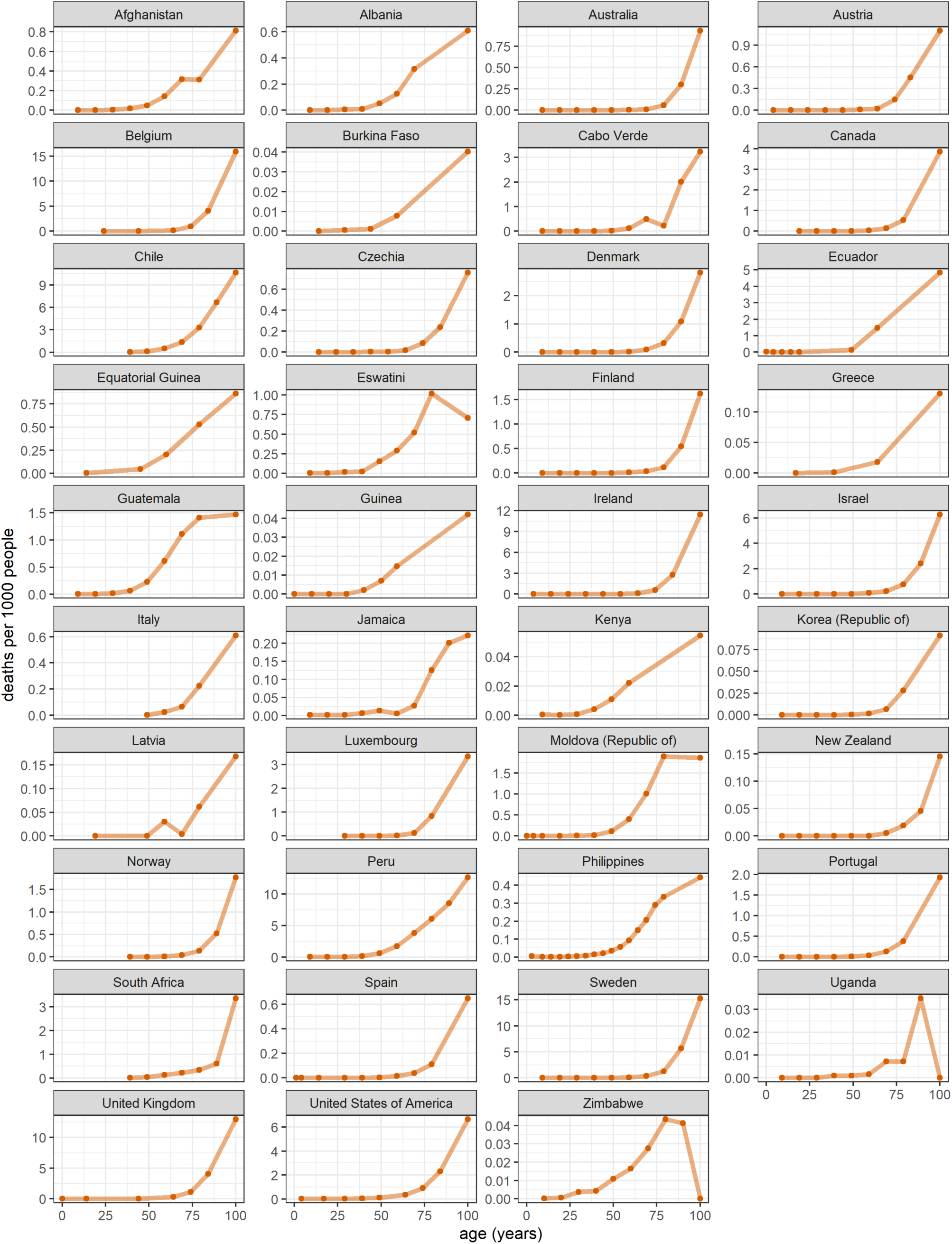
Age-specific cumulative death rate, by country or territory (observed deaths due to COVID-19 per 1000 population).

**Table S2.** Median and inter-quartile range of the age of cases and deaths, standardised by applying each country’s observed age-specific cumulative incidence and death rates to the world’s population age structure.

| Country or territory | Standardised median age of cases<br>(inter-quartile range) | Standardised median age of deaths<br>(inter-quartile range) |
| --- | --- | --- |
| Afghanistan | 47 (34 to 60) | 65 (55 to 76) |
| Albania | 41 (29 to 54) | 67 (58 to 82) |
| Australia | 32 (22 to 48) | 83 (75 to 89) |
| Austria | 30 (19 to 46) | 78 (70 to 87) |
| Belgium | 37 (24 to 54) | 81 (72 to 90) |
| Burkina Faso | 53 (38 to 73) | 73 (60 to 87) |
| Cabo Verde | 33 (22 to 51) | 69 (62 to 84) |
| Canada | 37 (24 to 53) | 84 (74 to 92) |
| Central African Republic | 40 (32 to 50) | n/a |
| Chile | 36 (25 to 51) | 72 (62 to 81) |
| Cyprus | 39 (26 to 53) | n/a |
| Czechia | 29 (18 to 44) | 78 (69 to 87) |
| Denmark | 33 (21 to 48) | 80 (71 to 87) |
| Ecuador | 49 (49 to 57) | 71 (57 to 85) |
| El Salvador | 44 (31 to 55) | n/a |
| Equatorial Guinea | 48 (32 to 56) | 63 (50 to 74) |
| Estonia | 37 (23 to 52) | n/a |
| Fiji | 34 (24 to 52) | n/a |
| Finland | 33 (22 to 47) | 80 (68 to 87) |
| Gambia | 45 (33 to 59) | n/a |
| Germany | 32 (20 to 48) | n/a |
| Greece | 32 (22 to 46) | 74 (59 to 87) |
| Guatemala | 42 (30 to 55) | 62 (53 to 72) |
| Guinea | 42 (31 to 55) | 71 (56 to 85) |
| Hong Kong | 33 (20 to 50) | n/a |
| Indonesia | 39 (27 to 53) | n/a |
| Ireland | 37 (26 to 54) | 82 (74 to 90) |
| Israel | 28 (17 to 45) | 78 (68 to 86) |
| Italy | n/a | 75 (66 to 86) |
| Jamaica | 35 (24 to 49) | 71 (50 to 79) |
| Kenya | 41 (31 to 54) | 67 (52 to 83) |
| Korea (Republic of) | 33 (22 to 53) | 78 (70 to 89) |
| Latvia | 34 (23 to 48) | 73 (56 to 85) |
| Lebanon | 39 (26 to 57) | n/a |
| Lithuania | 36 (24 to 49) | n/a |
| Luxembourg | 27 (16 to 42) | 82 (73 to 91) |
| Malawi | 40 (31 to 51) | n/a |
| Maldives | 35 (20 to 50) | n/a |
| Mexico | 45 (33 to 58) | n/a |
| Moldova (Republic of) | 44 (28 to 57) | 67 (59 to 75) |
| Montenegro | 35 (24 to 50) | n/a |
| Mozambique | 38 (29 to 52) | n/a |
| Nepal | 34 (25 to 44) | n/a |
| Netherlands | 33 (22 to 47) | n/a |
| New Zealand | 33 (23 to 49) | 78 (71 to 86) |
| North Macedonia | 39 (26 to 54) | n/a |
| Norway | 33 (22 to 48) | 80 (70 to 88) |
| Palestine (State of) | 41 (25 to 56) | n/a |
| Panama | 37 (25 to 52) | n/a |
| Paraguay | 38 (28 to 52) | n/a |
| Peru | n/a | 66 (57 to 76) |
| Philippines | 39 (28 to 54) | 66 (56 to 75) |
| Portugal | 33 (22 to 48) | 80 (69 to 90) |
| South Africa | n/a | 67 (55 to 80) |
| Spain | 28 (16 to 42) | 81 (68 to 90) |
| Swaziland | 42 (31 to 54) | 64 (53 to 73) |
| Sweden | 38 (26 to 53) | 81 (71 to 87) |
| Switzerland | 42 (31 to 55) | n/a |
| Uganda | 38 (30 to 47) | 68 (59 to 81) |
| United Kingdom | n/a | 77 (67 to 87) |
| United States of America | 35 (23 to 50) | 73 (60 to 83) |
| Venezuela (Bolivarian Republic of) | 33 (22 to 46) | n/a |
| Vietnam | 38 (25 to 55) | n/a |
| Zambia | 44 (33 to 65) | n/a |
| Zimbabwe | 41 (31 to 53) | 59 (46 to 70) |

**Table S3.**
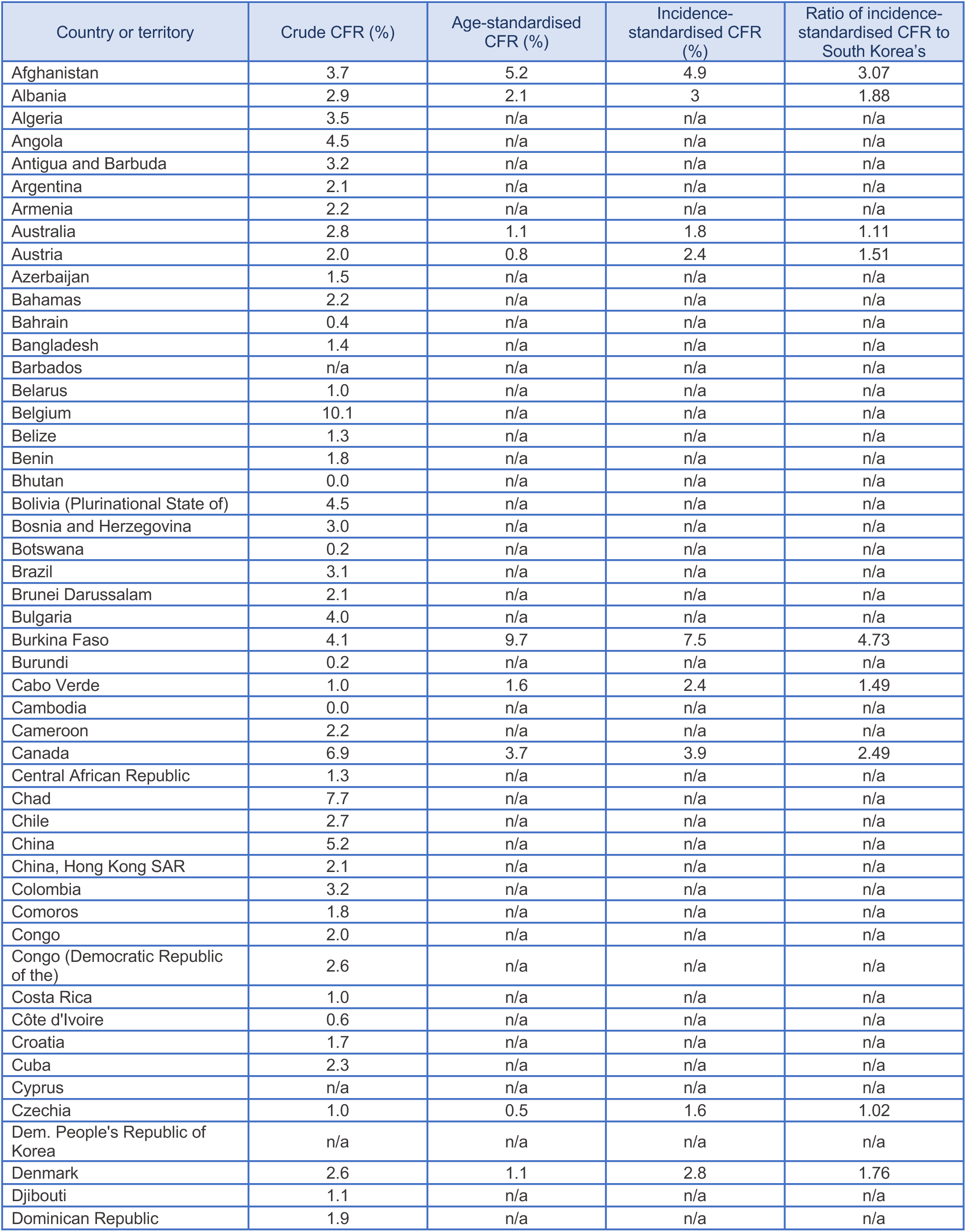
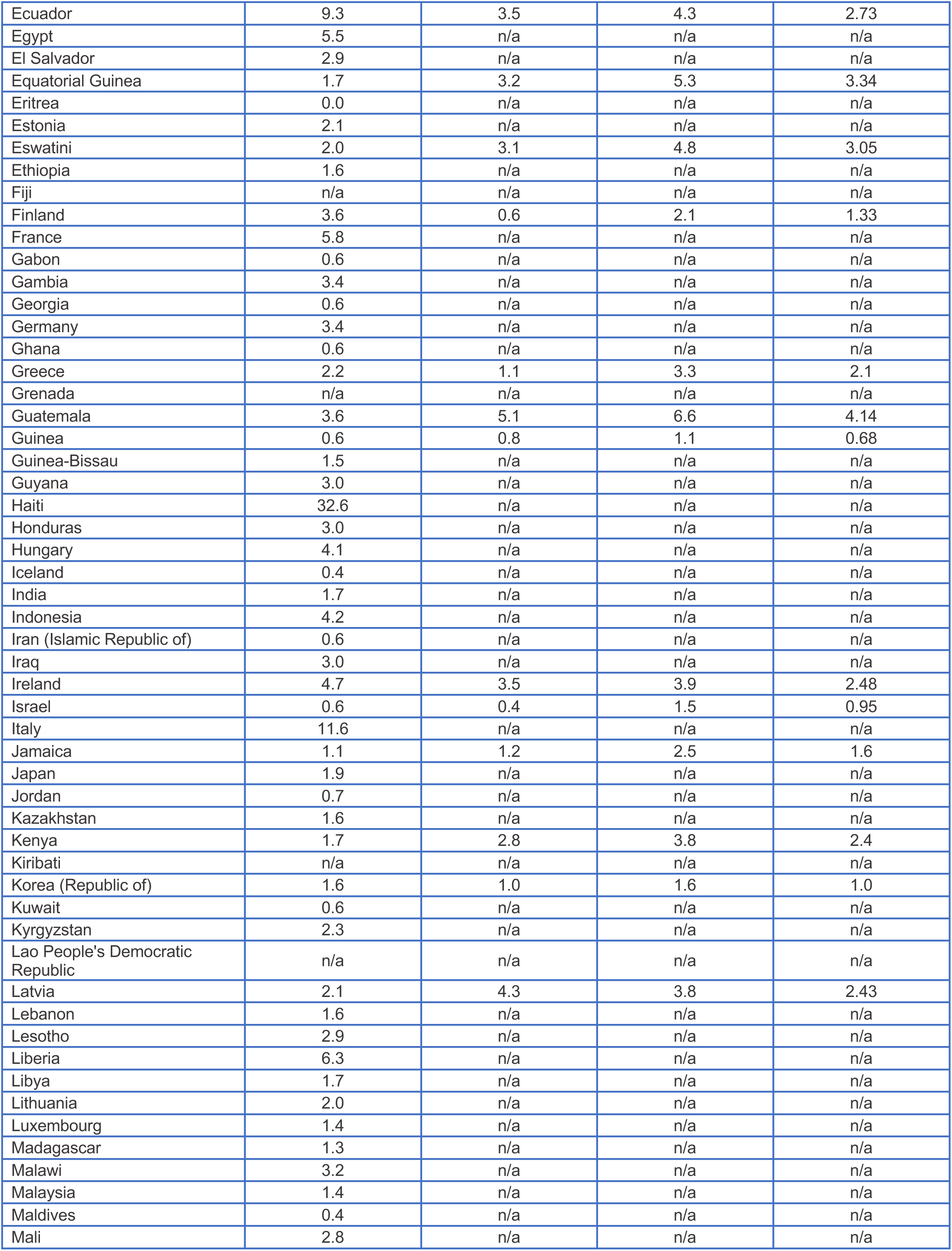
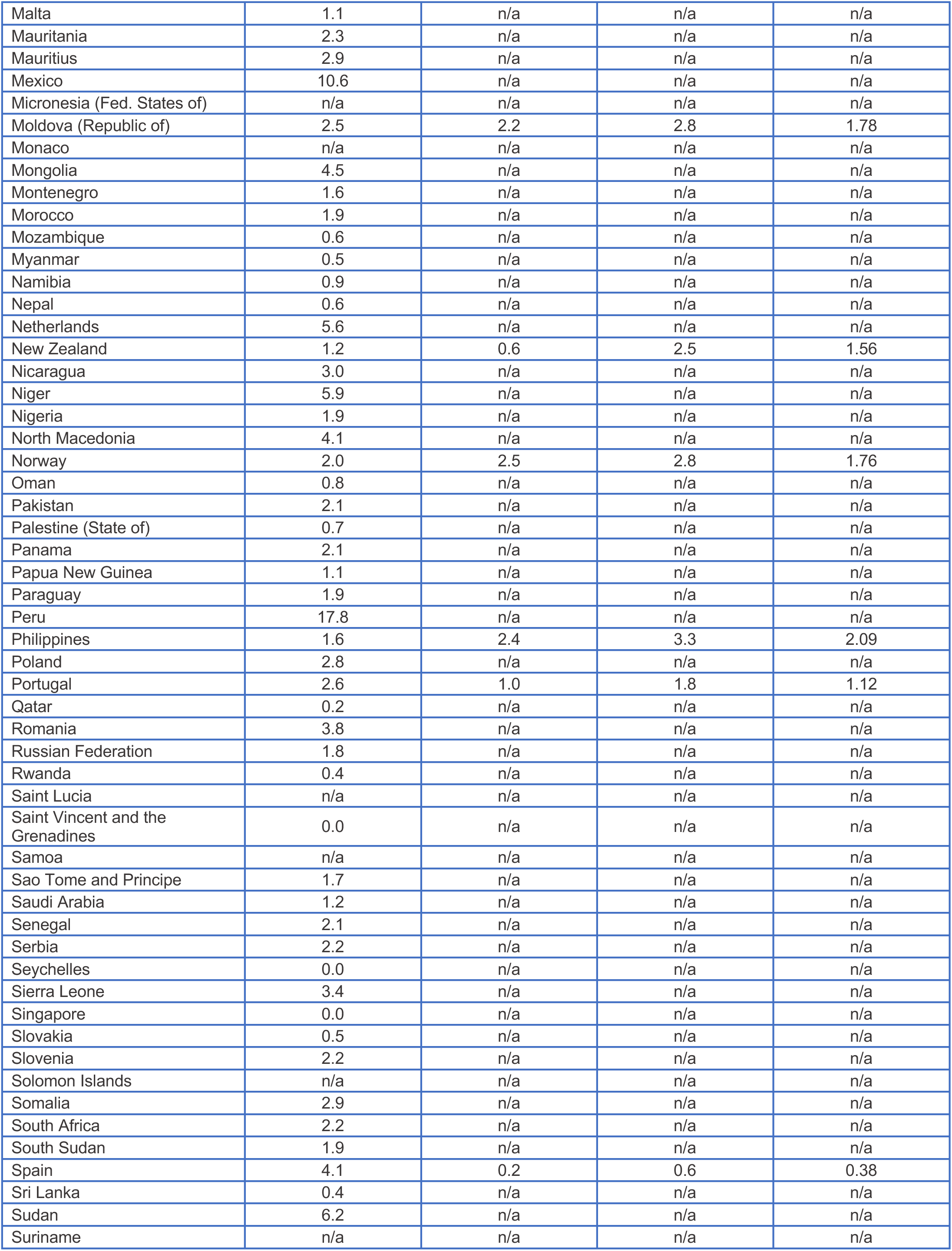
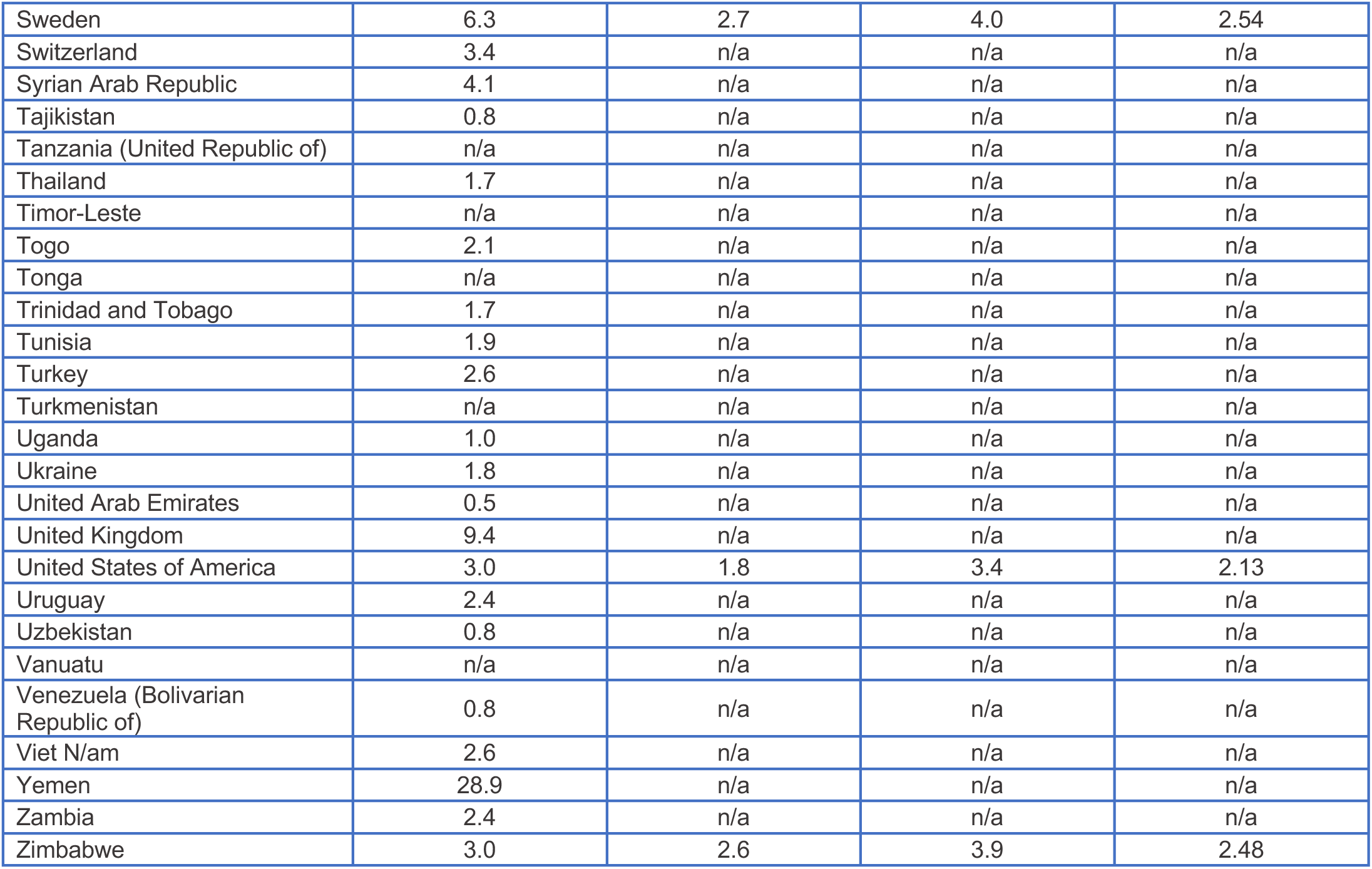
Crude, age-standardised and incidence-standardised case-fatality ratios, by country or territory. The last column is the ratio of incidence-standardised CFR for the country, relative to that of South Korea.

| Country or territory | Crude CFR (%) | Age-standardised CFR (%) | Incidence-standardised CFR (%) | Ratio of incidence-standardised CFR to South Korea's |
| --- | --- | --- | --- | --- |
| Afghanistan | 3.7 | 5.2 | 4.9 | 3.07 |
| Albania | 2.9 | 2.1 | 3 | 1.88 |
| Algeria | 3.5 | n/a | n/a | n/a |
| Angola | 4.5 | n/a | n/a | n/a |
| Antigua and Barbuda | 3.2 | n/a | n/a | n/a |
| Argentina | 2.1 | n/a | n/a | n/a |
| Armenia | 2.2 | n/a | n/a | n/a |
| Australia | 2.8 | 1.1 | 1.8 | 1.11 |
| Austria | 2.0 | 0.8 | 2.4 | 1.51 |
| Azerbaijan | 1.5 | n/a | n/a | n/a |
| Bahamas | 2.2 | n/a | n/a | n/a |
| Bahrain | 0.4 | n/a | n/a | n/a |
| Bangladesh | 1.4 | n/a | n/a | n/a |
| Barbados | n/a | n/a | n/a | n/a |
| Belarus | 1.0 | n/a | n/a | n/a |
| Belgium | 10.1 | n/a | n/a | n/a |
| Belize | 1.3 | n/a | n/a | n/a |
| Benin | 1.8 | n/a | n/a | n/a |
| Bhutan | 0.0 | n/a | n/a | n/a |
| Bolivia (Plurinational State of) | 4.5 | n/a | n/a | n/a |
| Bosnia and Herzegovina | 3.0 | n/a | n/a | n/a |
| Botswana | 0.2 | n/a | n/a | n/a |
| Brazil | 3.1 | n/a | n/a | n/a |
| Brunei Darussalam | 2.1 | n/a | n/a | n/a |
| Bulgaria | 4.0 | n/a | n/a | n/a |
| Burkina Faso | 4.1 | 9.7 | 7.5 | 4.73 |
| Burundi | 0.2 | n/a | n/a | n/a |
| Cabo Verde | 1.0 | 1.6 | 2.4 | 1.49 |
| Cambodia | 0.0 | n/a | n/a | n/a |
| Cameroon | 2.2 | n/a | n/a | n/a |
| Canada | 6.9 | 3.7 | 3.9 | 2.49 |
| Central African Republic | 1.3 | n/a | n/a | n/a |
| Chad | 7.7 | n/a | n/a | n/a |
| Chile | 2.7 | n/a | n/a | n/a |
| China | 5.2 | n/a | n/a | n/a |
| China, Hong Kong SAR | 2.1 | n/a | n/a | n/a |
| Colombia | 3.2 | n/a | n/a | n/a |
| Comoros | 1.8 | n/a | n/a | n/a |
| Congo | 2.0 | n/a | n/a | n/a |
| Congo (Democratic Republic of the) | 2.6 | n/a | n/a | n/a |
| Costa Rica | 1.0 | n/a | n/a | n/a |
| Côte d'Ivoire | 0.6 | n/a | n/a | n/a |
| Croatia | 1.7 | n/a | n/a | n/a |
| Cuba | 2.3 | n/a | n/a | n/a |
| Cyprus | n/a | n/a | n/a | n/a |
| Czechia | 1.0 | 0.5 | 1.6 | 1.02 |
| Dem. People's Republic of Korea | n/a | n/a | n/a | n/a |
| Denmark | 2.6 | 1.1 | 2.8 | 1.76 |
| Djibouti | 1.1 | n/a | n/a | n/a |
| Dominican Republic | 1.9 | n/a | n/a | n/a |
| Ecuador | 9.3 | 3.5 | 4.3 | 2.73 |
| Egypt | 5.5 | n/a | n/a | n/a |
| El Salvador | 2.9 | n/a | n/a | n/a |
| Equatorial Guinea | 1.7 | 3.2 | 5.3 | 3.34 |
| Eritrea | 0.0 | n/a | n/a | n/a |
| Estonia | 2.1 | n/a | n/a | n/a |
| Eswatini | 2.0 | 3.1 | 4.8 | 3.05 |
| Ethiopia | 1.6 | n/a | n/a | n/a |
| Fiji | n/a | n/a | n/a | n/a |
| Finland | 3.6 | 0.6 | 2.1 | 1.33 |
| France | 5.8 | n/a | n/a | n/a |
| Gabon | 0.6 | n/a | n/a | n/a |
| Gambia | 3.4 | n/a | n/a | n/a |
| Georgia | 0.6 | n/a | n/a | n/a |
| Germany | 3.4 | n/a | n/a | n/a |
| Ghana | 0.6 | n/a | n/a | n/a |
| Greece | 2.2 | 1.1 | 3.3 | 2.1 |
| Grenada | n/a | n/a | n/a | n/a |
| Guatemala | 3.6 | 5.1 | 6.6 | 4.14 |
| Guinea | 0.6 | 0.8 | 1.1 | 0.68 |
| Guinea-Bissau | 1.5 | n/a | n/a | n/a |
| Guyana | 3.0 | n/a | n/a | n/a |
| Haiti | 32.6 | n/a | n/a | n/a |
| Honduras | 3.0 | n/a | n/a | n/a |
| Hungary | 4.1 | n/a | n/a | n/a |
| Iceland | 0.4 | n/a | n/a | n/a |
| India | 1.7 | n/a | n/a | n/a |
| Indonesia | 4.2 | n/a | n/a | n/a |
| Iran (Islamic Republic of) | 0.6 | n/a | n/a | n/a |
| Iraq | 3.0 | n/a | n/a | n/a |
| Ireland | 4.7 | 3.5 | 3.9 | 2.48 |
| Israel | 0.6 | 0.4 | 1.5 | 0.95 |
| Italy | 11.6 | n/a | n/a | n/a |
| Jamaica | 1.1 | 1.2 | 2.5 | 1.6 |
| Japan | 1.9 | n/a | n/a | n/a |
| Jordan | 0.7 | n/a | n/a | n/a |
| Kazakhstan | 1.6 | n/a | n/a | n/a |
| Kenya | 1.7 | 2.8 | 3.8 | 2.4 |
| Kiribati | n/a | n/a | n/a | n/a |
| Korea (Republic of) | 1.6 | 1.0 | 1.6 | 1.0 |
| Kuwait | 0.6 | n/a | n/a | n/a |
| Kyrgyzstan | 2.3 | n/a | n/a | n/a |
| Lao People's Democratic Republic | n/a | n/a | n/a | n/a |
| Latvia | 2.1 | 4.3 | 3.8 | 2.43 |
| Lebanon | 1.6 | n/a | n/a | n/a |
| Lesotho | 2.9 | n/a | n/a | n/a |
| Liberia | 6.3 | n/a | n/a | n/a |
| Libya | 1.7 | n/a | n/a | n/a |
| Lithuania | 2.0 | n/a | n/a | n/a |
| Luxembourg | 1.4 | n/a | n/a | n/a |
| Madagascar | 1.3 | n/a | n/a | n/a |
| Malawi | 3.2 | n/a | n/a | n/a |
| Malaysia | 1.4 | n/a | n/a | n/a |
| Maldives | 0.4 | n/a | n/a | n/a |
| Mali | 2.8 | n/a | n/a | n/a |
| Malta | 1.1 | n/a | n/a | n/a |
| Mauritania | 2.3 | n/a | n/a | n/a |
| Mauritius | 2.9 | n/a | n/a | n/a |
| Mexico | 10.6 | n/a | n/a | n/a |
| Micronesia (Fed. States of) | n/a | n/a | n/a | n/a |
| Moldova (Republic of) | 2.5 | 2.2 | 2.8 | 1.78 |
| Monaco | n/a | n/a | n/a | n/a |
| Mongolia | 4.5 | n/a | n/a | n/a |
| Montenegro | 1.6 | n/a | n/a | n/a |
| Morocco | 1.9 | n/a | n/a | n/a |
| Mozambique | 0.6 | n/a | n/a | n/a |
| Myanmar | 0.5 | n/a | n/a | n/a |
| Namibia | 0.9 | n/a | n/a | n/a |
| Nepal | 0.6 | n/a | n/a | n/a |
| Netherlands | 5.6 | n/a | n/a | n/a |
| New Zealand | 1.2 | 0.6 | 2.5 | 1.56 |
| Nicaragua | 3.0 | n/a | n/a | n/a |
| Niger | 5.9 | n/a | n/a | n/a |
| Nigeria | 1.9 | n/a | n/a | n/a |
| North Macedonia | 4.1 | n/a | n/a | n/a |
| Norway | 2.0 | 2.5 | 2.8 | 1.76 |
| Oman | 0.8 | n/a | n/a | n/a |
| Pakistan | 2.1 | n/a | n/a | n/a |
| Palestine (State of) | 0.7 | n/a | n/a | n/a |
| Panama | 2.1 | n/a | n/a | n/a |
| Papua New Guinea | 1.1 | n/a | n/a | n/a |
| Paraguay | 1.9 | n/a | n/a | n/a |
| Peru | 17.8 | n/a | n/a | n/a |
| Philippines | 1.6 | 2.4 | 3.3 | 2.09 |
| Poland | 2.8 | n/a | n/a | n/a |
| Portugal | 2.6 | 1.0 | 1.8 | 1.12 |
| Qatar | 0.2 | n/a | n/a | n/a |
| Romania | 3.8 | n/a | n/a | n/a |
| Russian Federation | 1.8 | n/a | n/a | n/a |
| Rwanda | 0.4 | n/a | n/a | n/a |
| Saint Lucia | n/a | n/a | n/a | n/a |
| Saint Vincent and the Grenadines | 0.0 | n/a | n/a | n/a |
| Samoa | n/a | n/a | n/a | n/a |
| Sao Tome and Principe | 1.7 | n/a | n/a | n/a |
| Saudi Arabia | 1.2 | n/a | n/a | n/a |
| Senegal | 2.1 | n/a | n/a | n/a |
| Serbia | 2.2 | n/a | n/a | n/a |
| Seychelles | 0.0 | n/a | n/a | n/a |
| Sierra Leone | 3.4 | n/a | n/a | n/a |
| Singapore | 0.0 | n/a | n/a | n/a |
| Slovakia | 0.5 | n/a | n/a | n/a |
| Slovenia | 2.2 | n/a | n/a | n/a |
| Solomon Islands | n/a | n/a | n/a | n/a |
| Somalia | 2.9 | n/a | n/a | n/a |
| South Africa | 2.2 | n/a | n/a | n/a |
| South Sudan | 1.9 | n/a | n/a | n/a |
| Spain | 4.1 | 0.2 | 0.6 | 0.38 |
| Sri Lanka | 0.4 | n/a | n/a | n/a |
| Sudan | 6.2 | n/a | n/a | n/a |
| Suriname | n/a | n/a | n/a | n/a |
| THIS ARTICLE HAS NOT YET BEEN PEER-REVIEWED |  |  |  |  |
| Sweden | 6.3 | 2.7 | 4.0 | 2.54 |
| Switzerland | 3.4 | n/a | n/a | n/a |
| Syrian Arab Republic | 4.1 | n/a | n/a | n/a |
| Tajikistan | 0.8 | n/a | n/a | n/a |
| Tanzania (United Republic of) | n/a | n/a | n/a | n/a |
| Thailand | 1.7 | n/a | n/a | n/a |
| Timor-Leste | n/a | n/a | n/a | n/a |
| Togo | 2.1 | n/a | n/a | n/a |
| Tonga | n/a | n/a | n/a | n/a |
| Trinidad and Tobago | 1.7 | n/a | n/a | n/a |
| Tunisia | 1.9 | n/a | n/a | n/a |
| Turkey | 2.6 | n/a | n/a | n/a |
| Turkmenistan | n/a | n/a | n/a | n/a |
| Uganda | 1.0 | n/a | n/a | n/a |
| Ukraine | 1.8 | n/a | n/a | n/a |
| United Arab Emirates | 0.5 | n/a | n/a | n/a |
| United Kingdom | 9.4 | n/a | n/a | n/a |
| United States of America | 3.0 | 1.8 | 3.4 | 2.13 |
| Uruguay | 2.4 | n/a | n/a | n/a |
| Uzbekistan | 0.8 | n/a | n/a | n/a |
| Vanuatu | n/a | n/a | n/a | n/a |
| Venezuela (Bolivarian Republic of) | 0.8 | n/a | n/a | n/a |
| Viet N/am | 2.6 | n/a | n/a | n/a |
| Yemen | 28.9 | n/a | n/a | n/a |
| Zambia | 2.4 | n/a | n/a | n/a |
| Zimbabwe | 3.0 | 2.6 | 3.9 | 2.48 |

**Figure S3.**
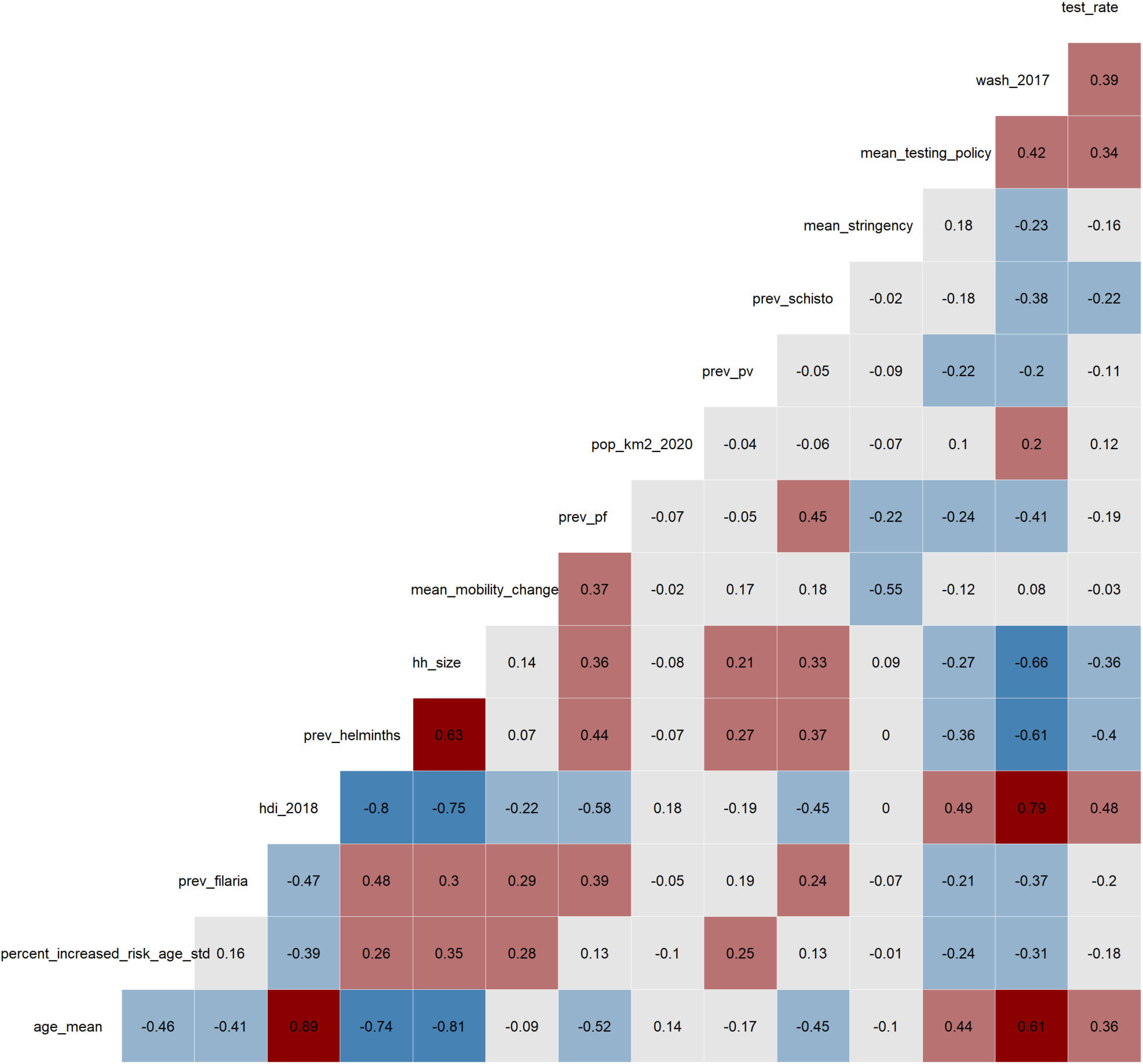
Variable collinearity heatmap. Values in each cell are the Pearson correlation coefficient for a given pair of variables. Coefficients close to −1 or +1 indicate high correlation, with 0 indicating minimal correlation.

**Table S4.** Pairs of highly collinear variables (Pearson correlation coefficient > |0.7|).

| Variable 1 | Variable 2 |
| --- | --- |
| Mean age of the country's population | Human Development Index |
| Mean age of the country's population | Prevalence of helminths |
| Mean age of the country's population | Household size |
| Human Development Index | Prevalence of helminths |
| Human Development Index | Household size |
| Human Development Index | Sanitation index |

## Univariate correlations between the independent variables and outcomes

**Figure S4.**
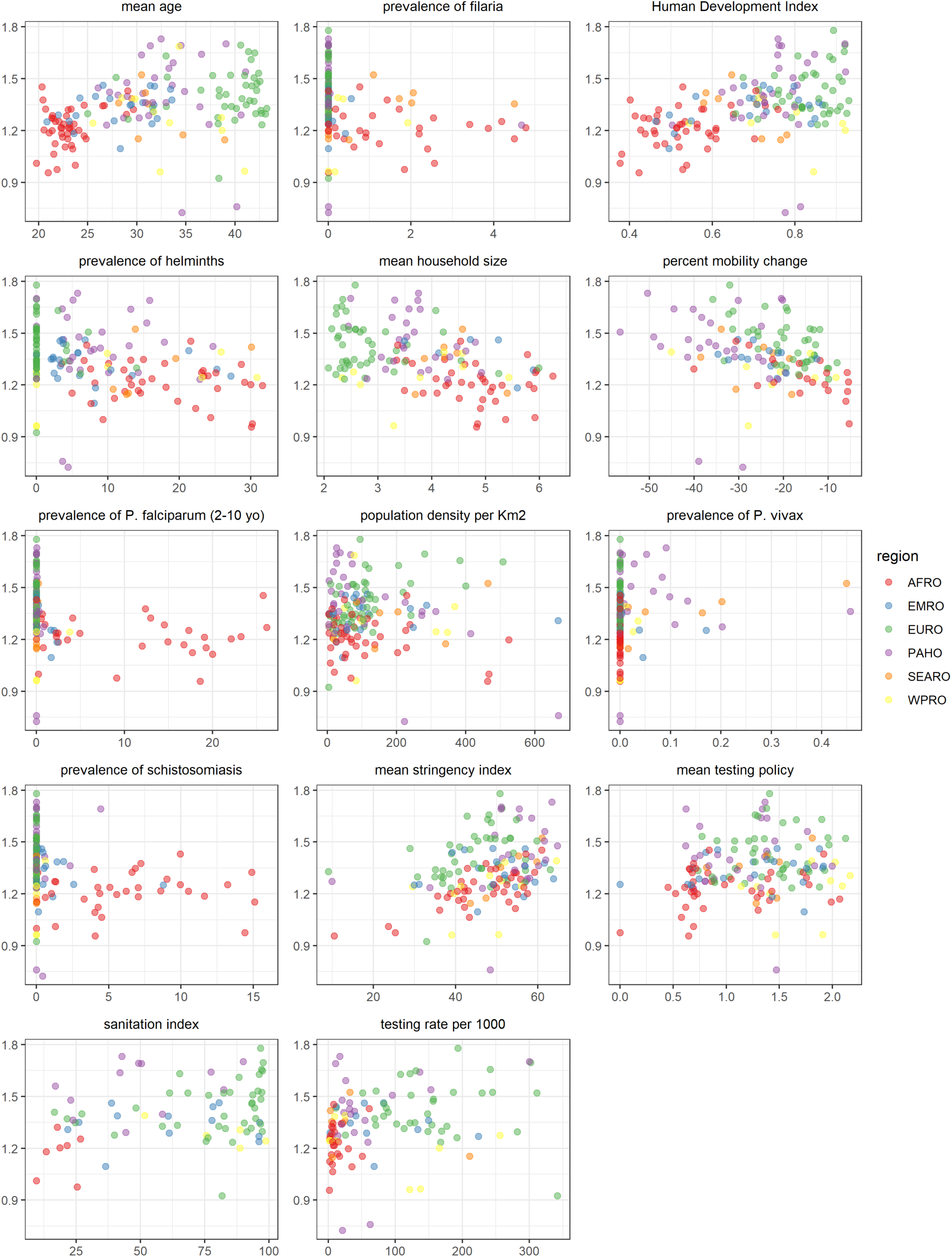
Correlation between each independent variable and the mean time-varying reproduction number, by WHO region. AFRO= African regional office; EMRO= Eastern Mediterranean regional office; EURO= European regional office; PAHO= Pan American health organisation; SEARO= South-East Asia regional office; WPRO= Western Pacific regional office

**Figure S5.**
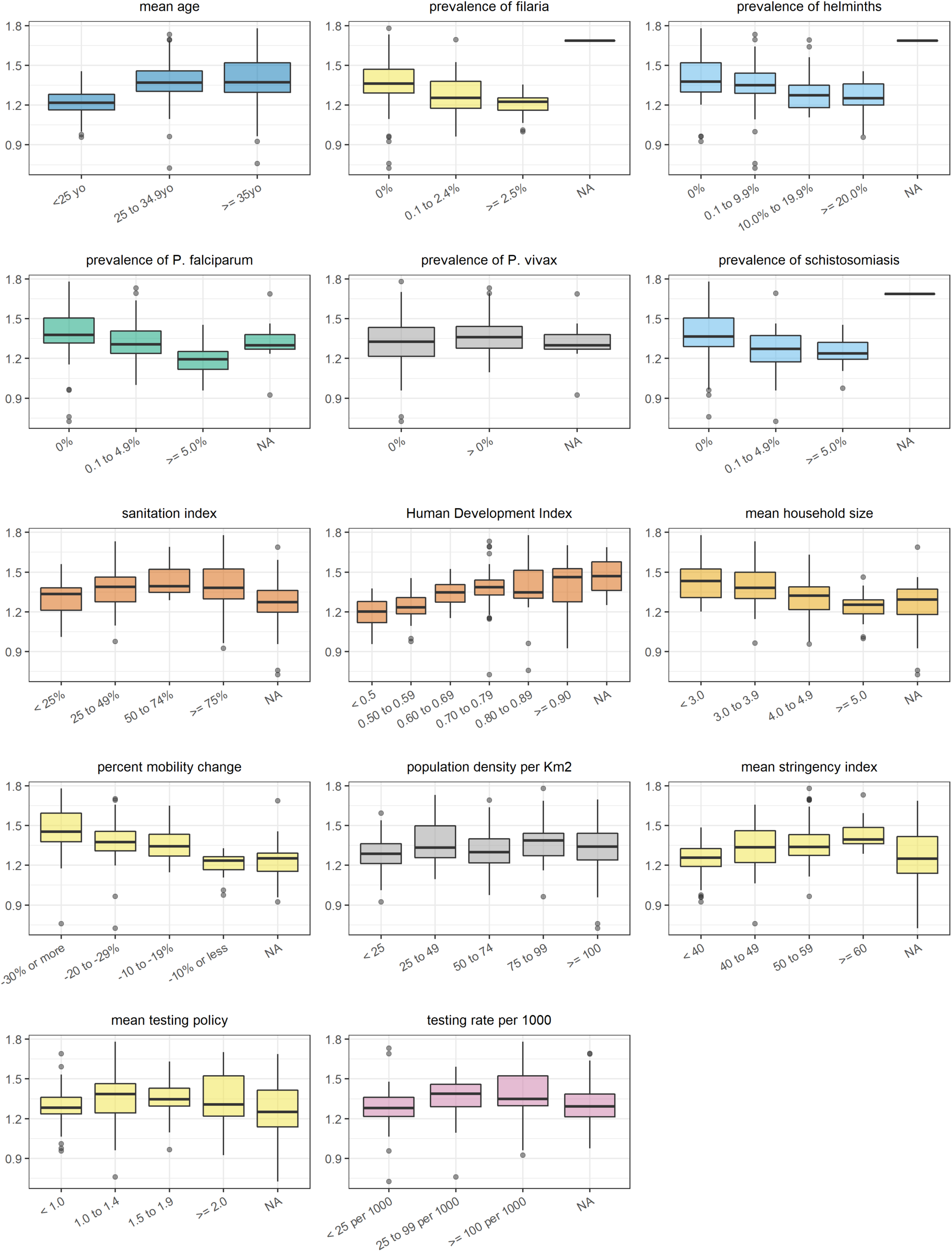
Correlation between each independent variable (categorised) and the mean time-varying reproduction number. Note: NA refers to countries for which no data were available for the specific variable considered

**Figure S6.**
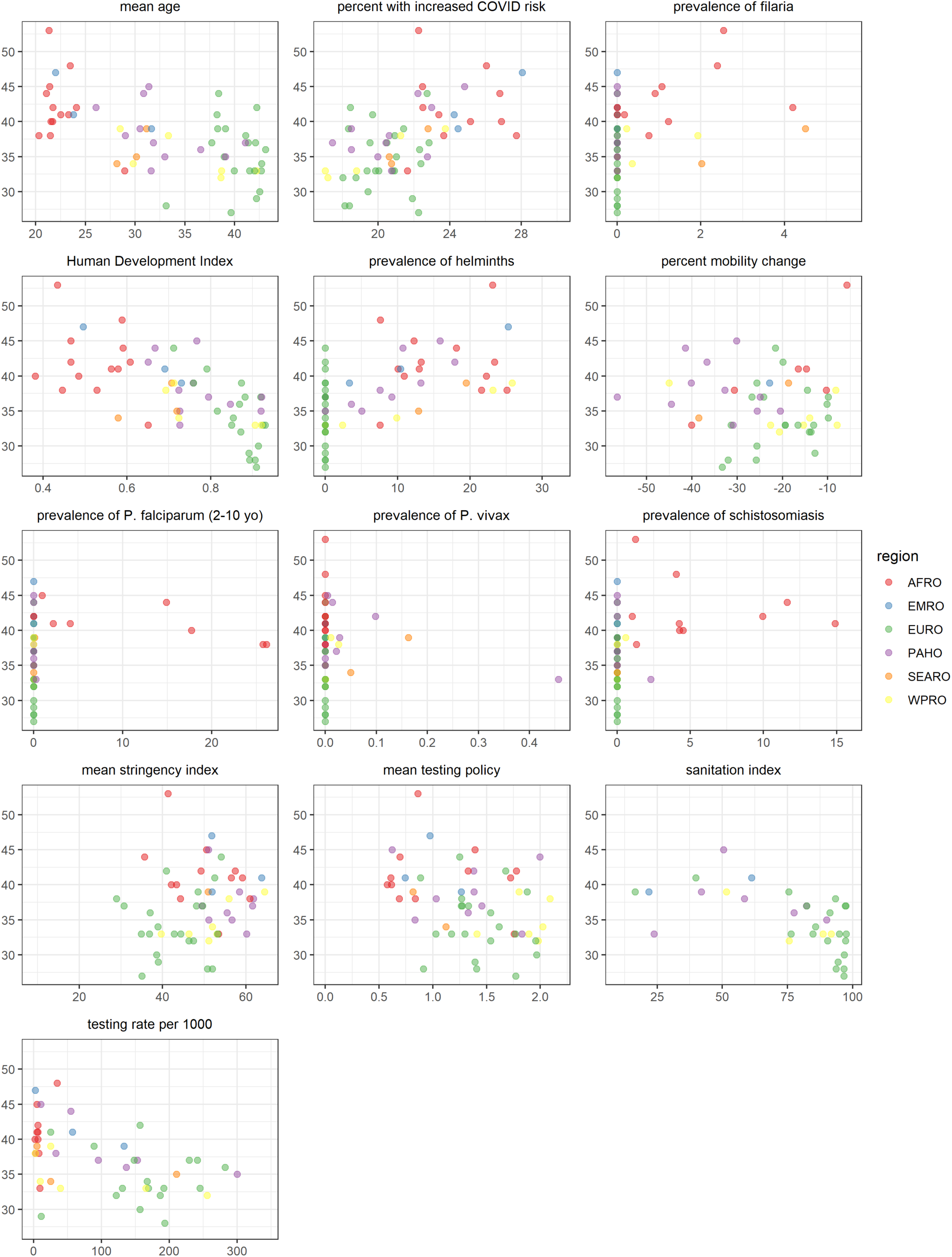
Correlation between each independent variable and age-standardised median age of cases, by WHO region. AFRO= African regional office; EMRO= Eastern Mediterranean regional office; EURO= European regional office; PAHO= Pan American health organisation; SEARO= South-East Asia regional office; WPRO= Western Pacific regional office

**Figure S7.**
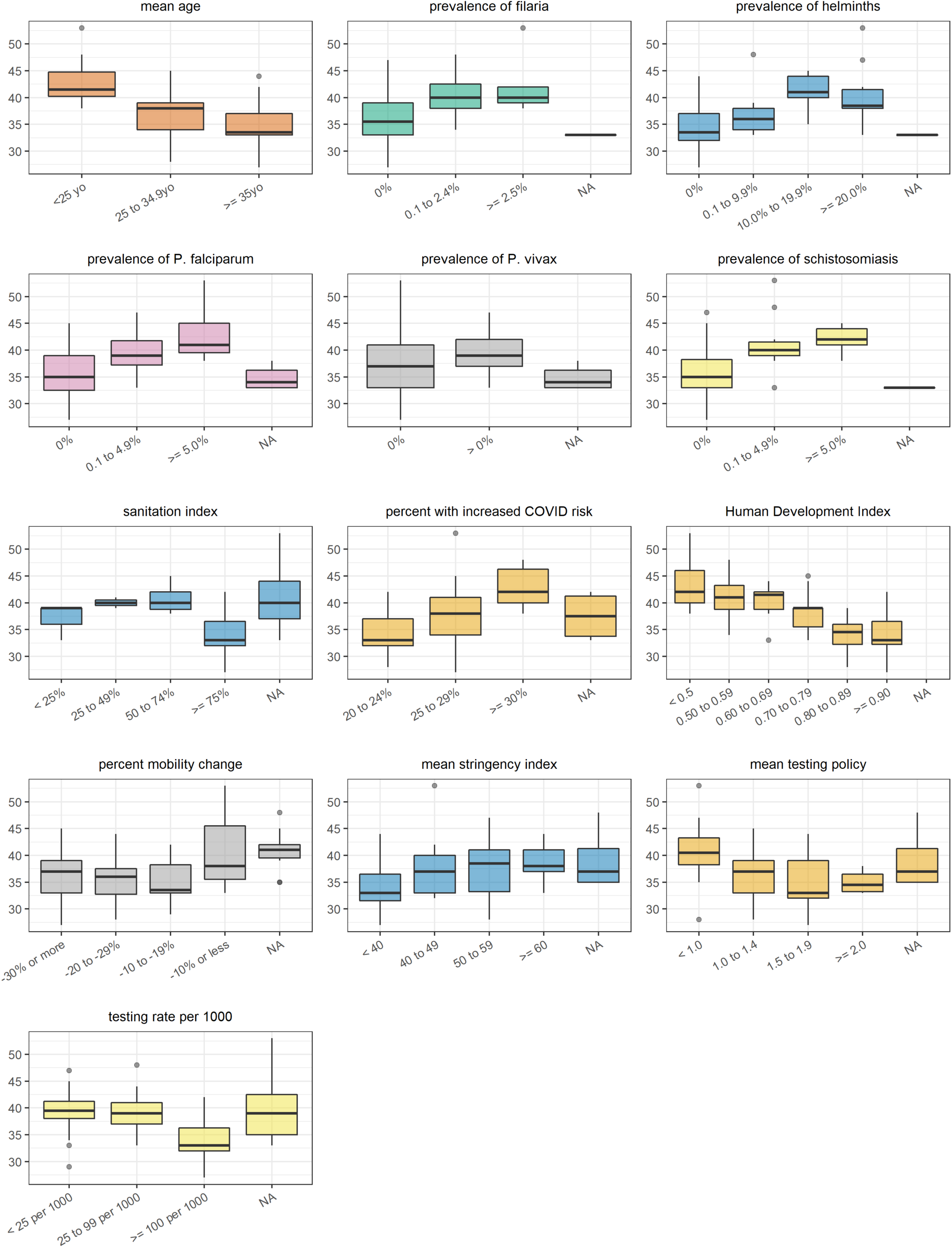
Correlation between each independent variable (categorised) and the age-standardised median age of cases. Note: NA refers to countries for which no data were available for the specific variable considered

**Figure S8.**
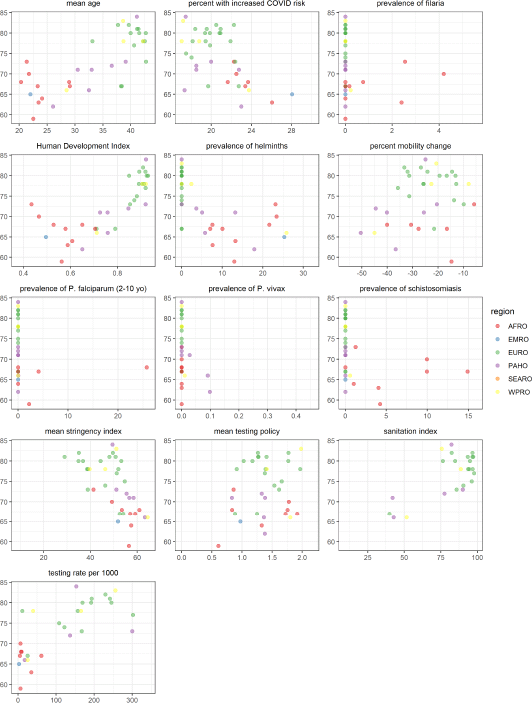
Correlation between each independent variable and age-standardised median age of deaths, by WHO region. AFRO= African regional office; EMRO= Eastern Mediterranean regional office; EURO= European regional office; PAHO= Pan American health organisation; SEARO= South-East Asia regional office; WPRO= Western Pacific regional office

**Figure S9.**
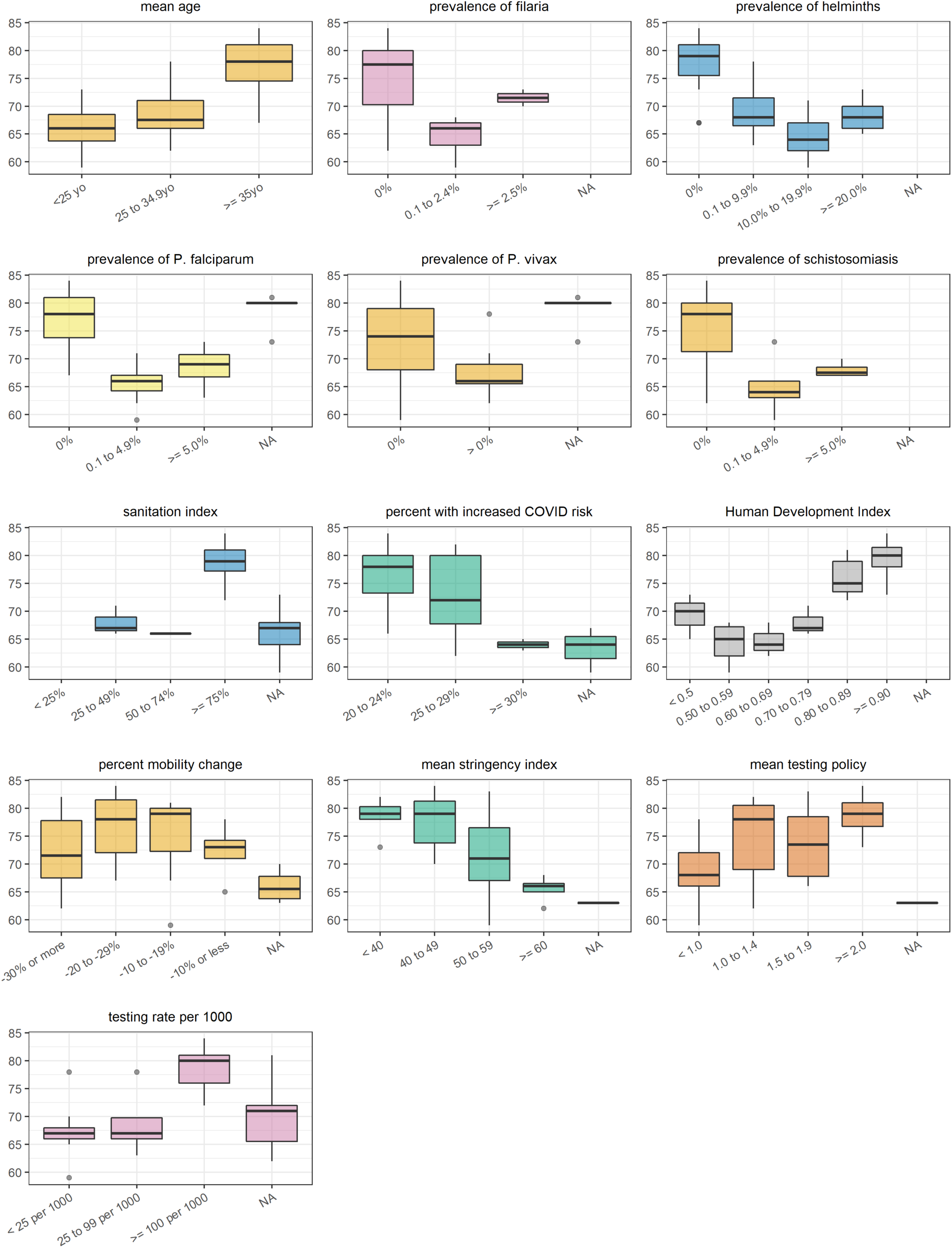
Correlation between each independent variable (categorised) and the age-standardised median age of deaths. Note: NA refers to countries for which no data were available for the specific variable considered

**Figure S10.**
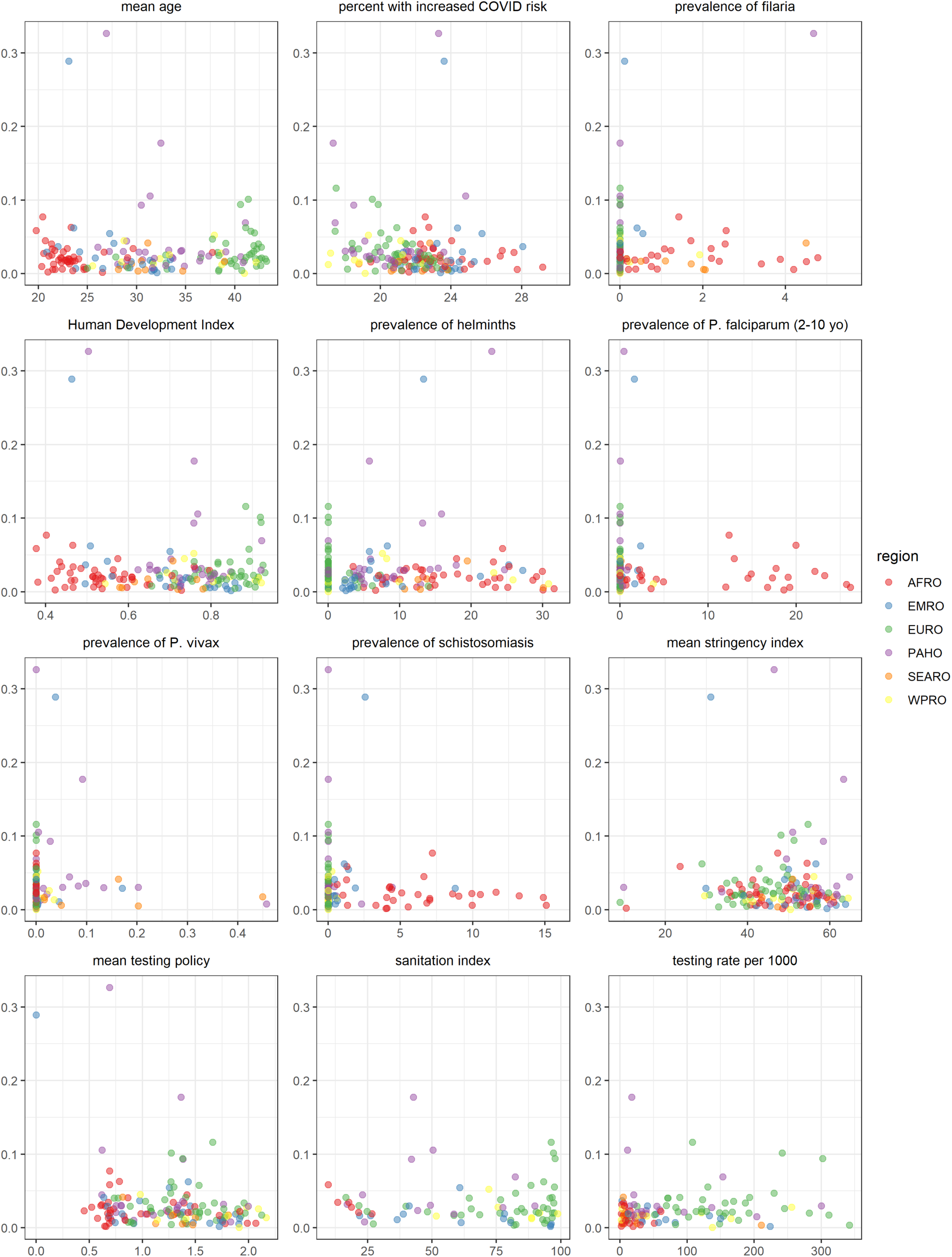
Correlation between each independent variable and crude case-fatality ratio, by WHO region. AFRO= African regional office; EMRO= Eastern Mediterranean regional office; EURO= European regional office; PAHO= Pan American health organisation; SEARO= South-East Asia regional office; WPRO= Western Pacific regional office

**Figure S11.**
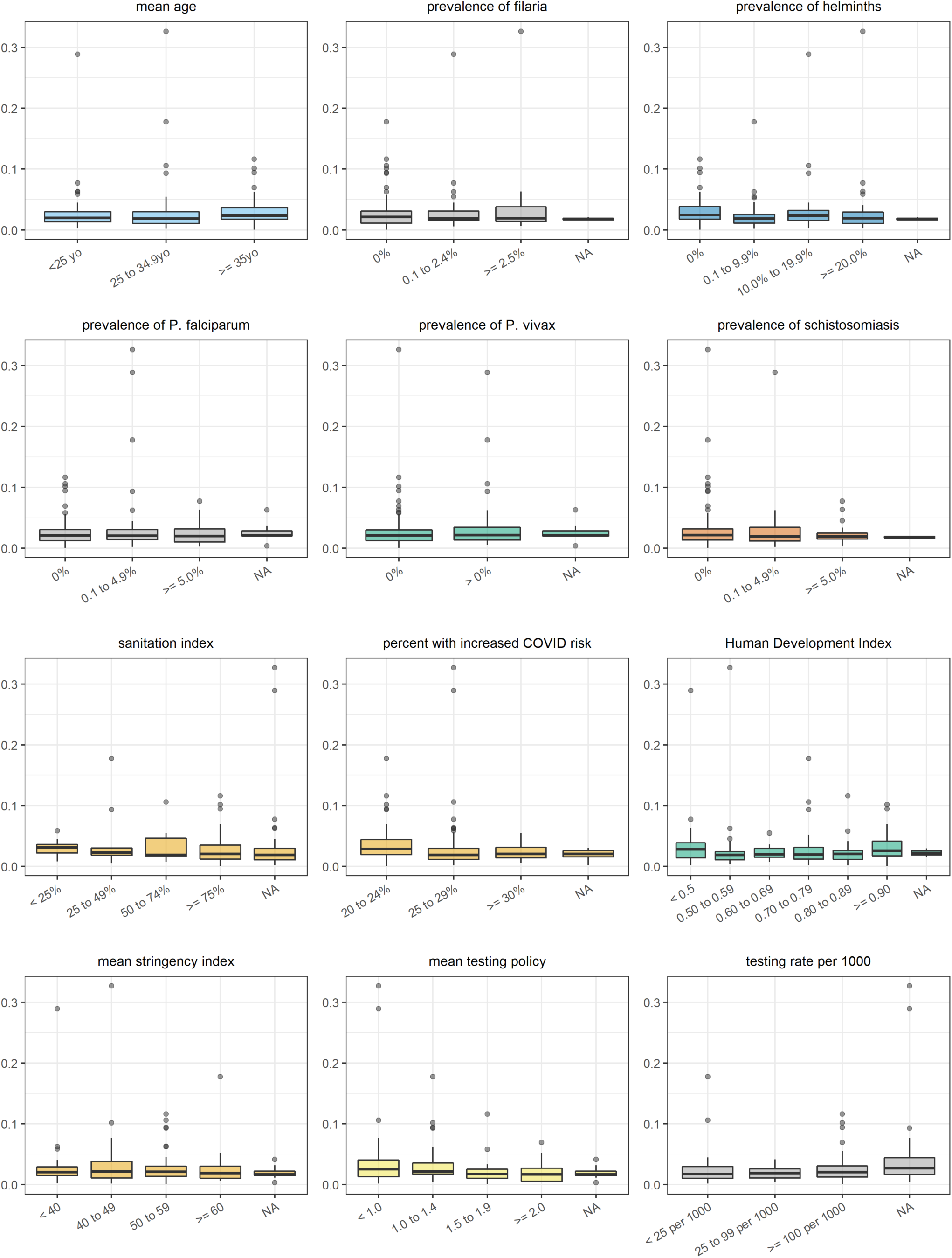
Correlation between each independent variable (categorised) and the crude case-fatality ratio. Note: NA refers to countries for which no data were available for the specific variable considered

**Figure S12.**
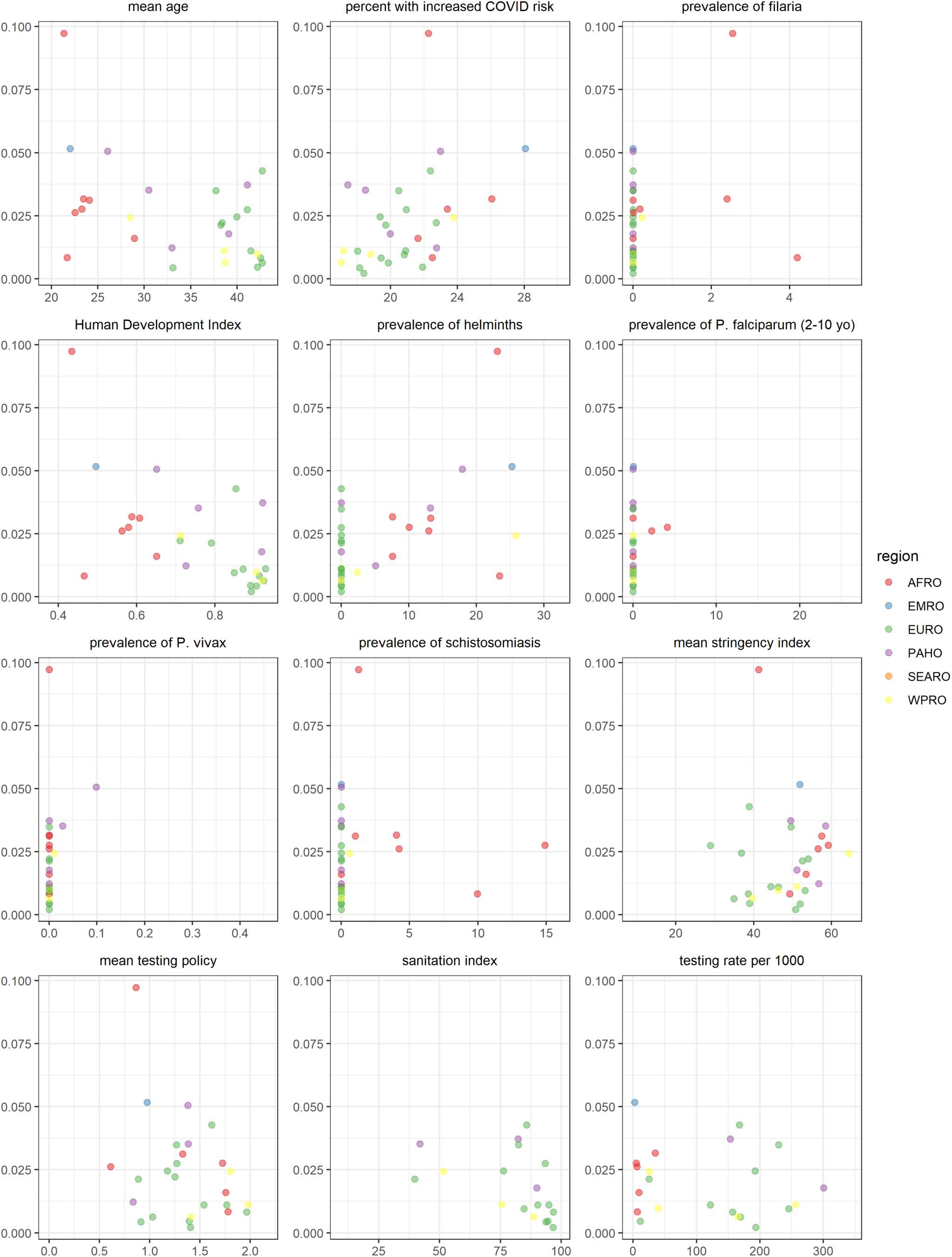
Correlation between each independent variable and age-standardised case-fatality ratio, by WHO region. AFRO= African regional office; EMRO= Eastern Mediterranean regional office; EURO= European regional office; PAHO= Pan American health organisation; SEARO= South-East Asia regional office; WPRO= Western Pacific regional office

**Figure S13.**
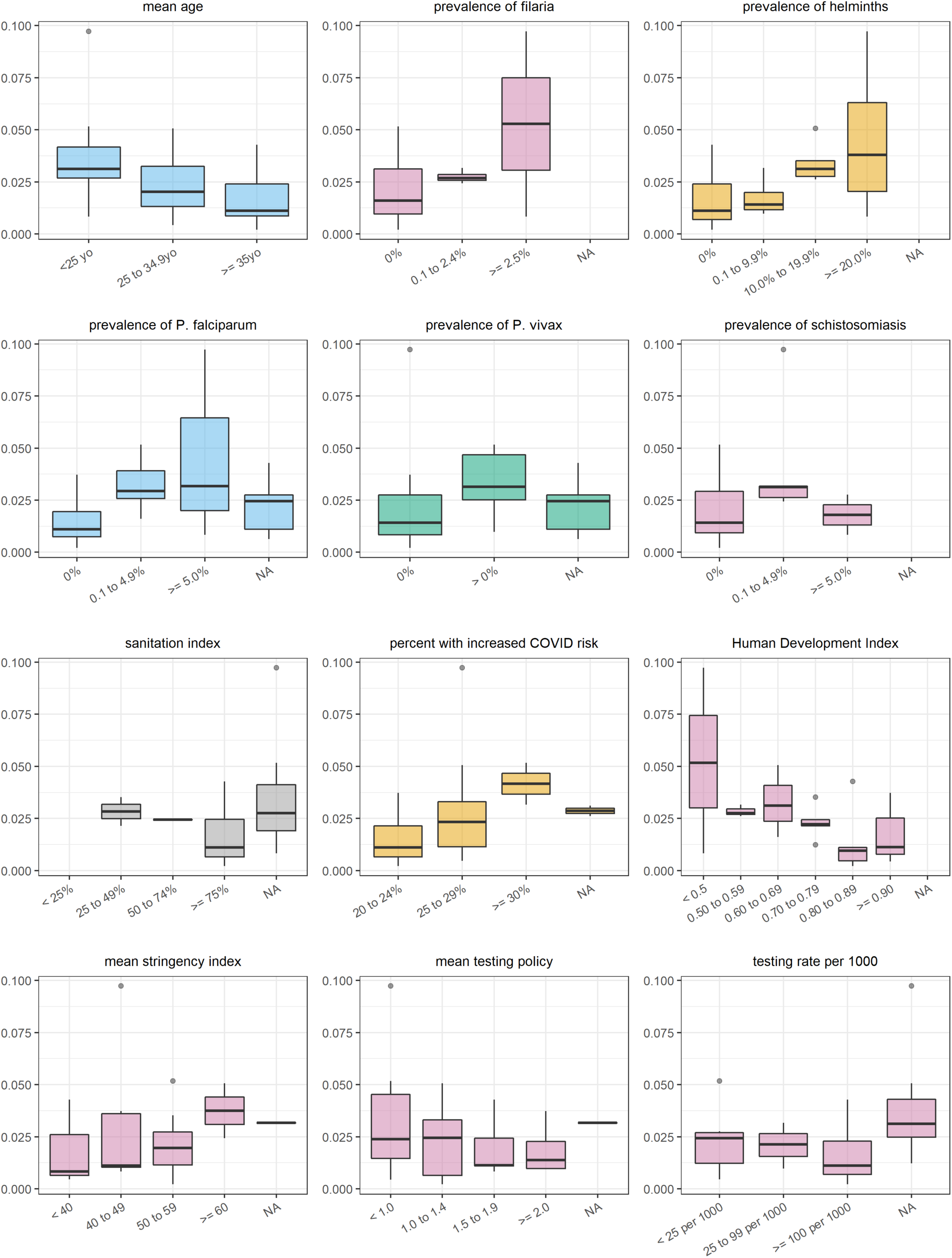
Correlation between each independent variable (categorised) and the age-standardised case-fatality ratio. Note: NA refers to countries for which no data were available for the specific variable considered

**Figure S14.**
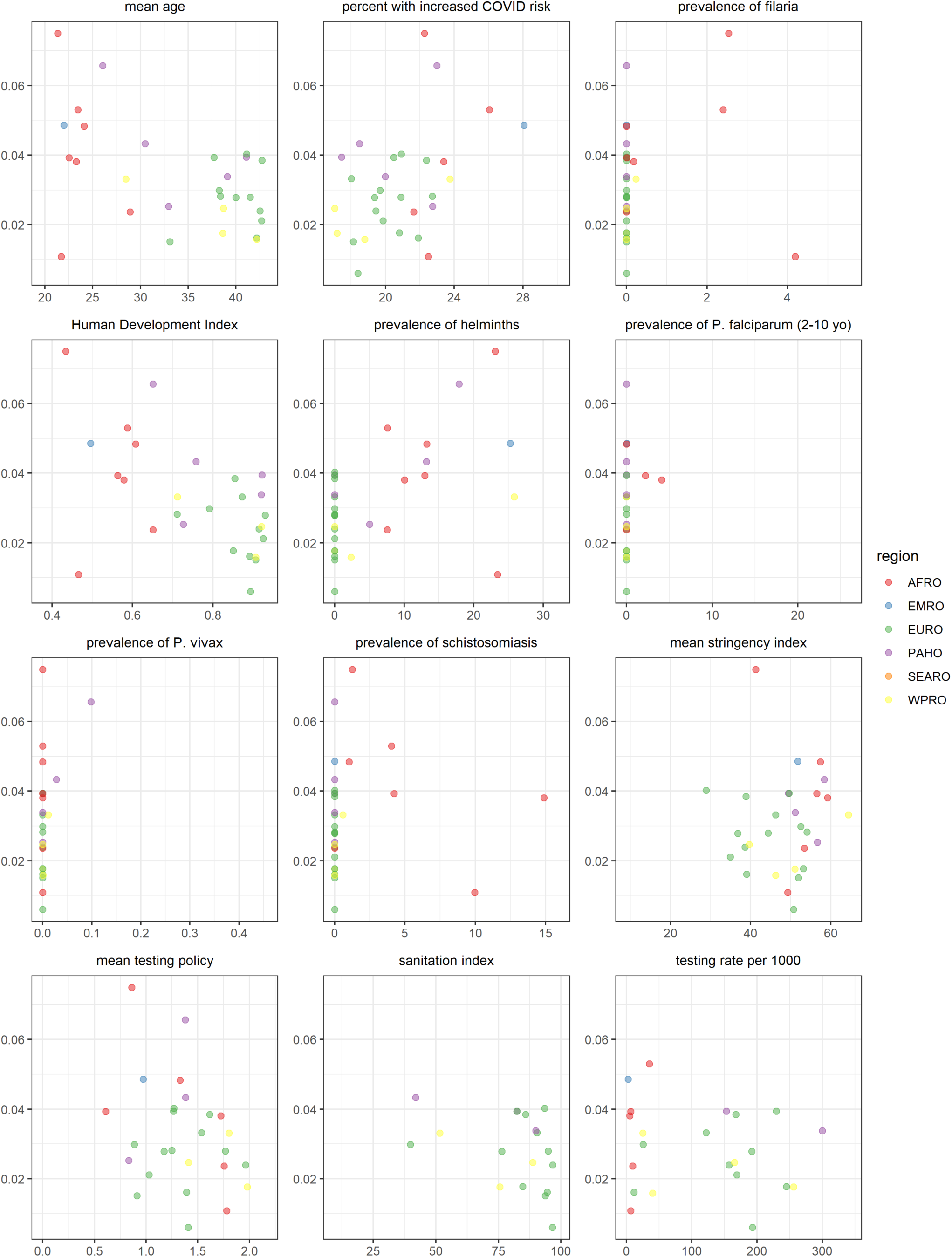
Correlation between each independent variable and incidence-standardised case-fatality ratio, by WHO region. AFRO= African regional office; EMRO= Eastern Mediterranean regional office; EURO= European regional office; PAHO= Pan American health organisation; SEARO= South-East Asia regional office; WPRO= Western Pacific regional office

**Figure S15.**
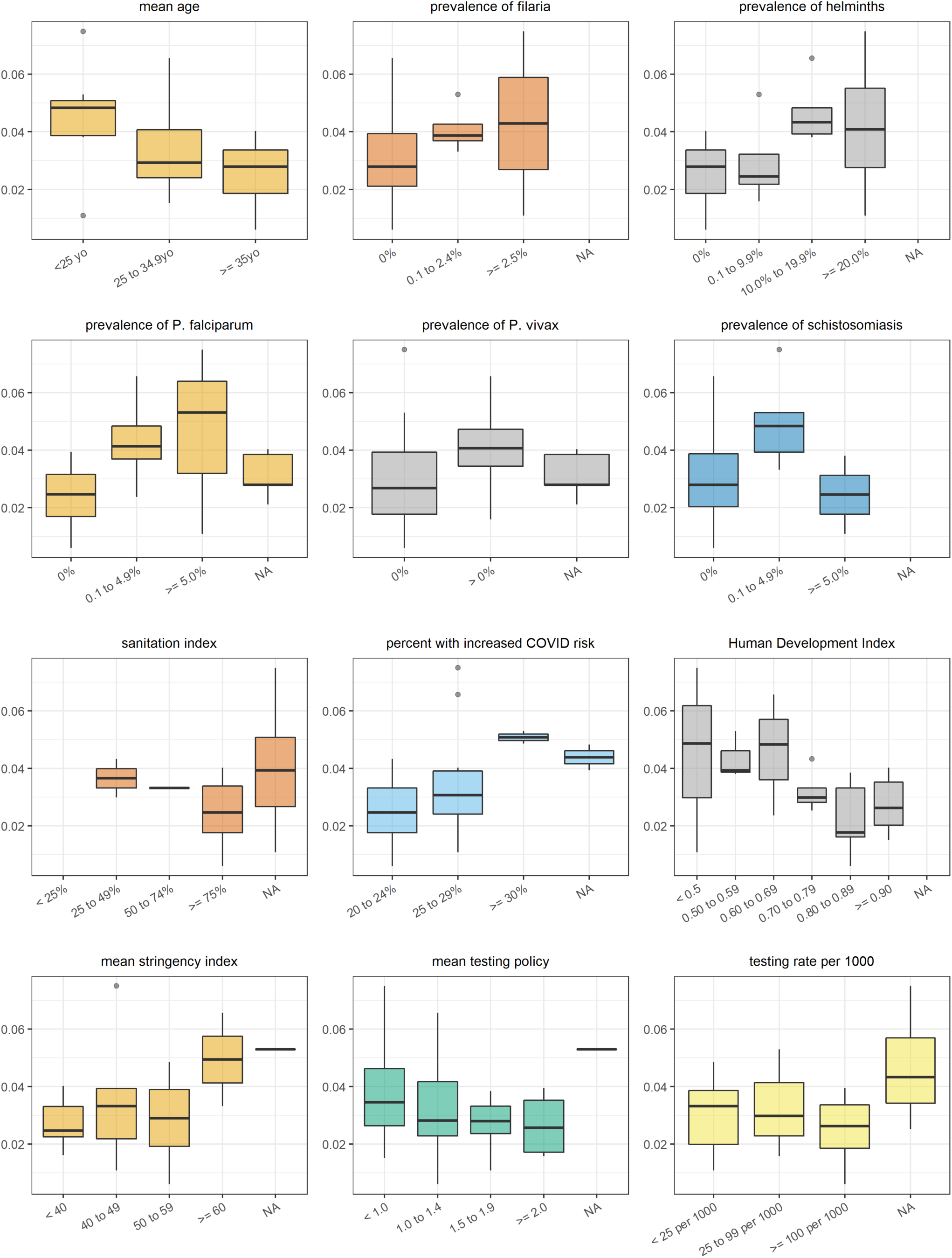
Correlation between each independent variable (categorised) and the incidence-standardised case-fatality ratio. Note: NA refers to countries for which no data were available for the specific variable considered

## Detailed multivariate analysis results

Note: all results are presented for the models imputing missing data

**Table S4.**
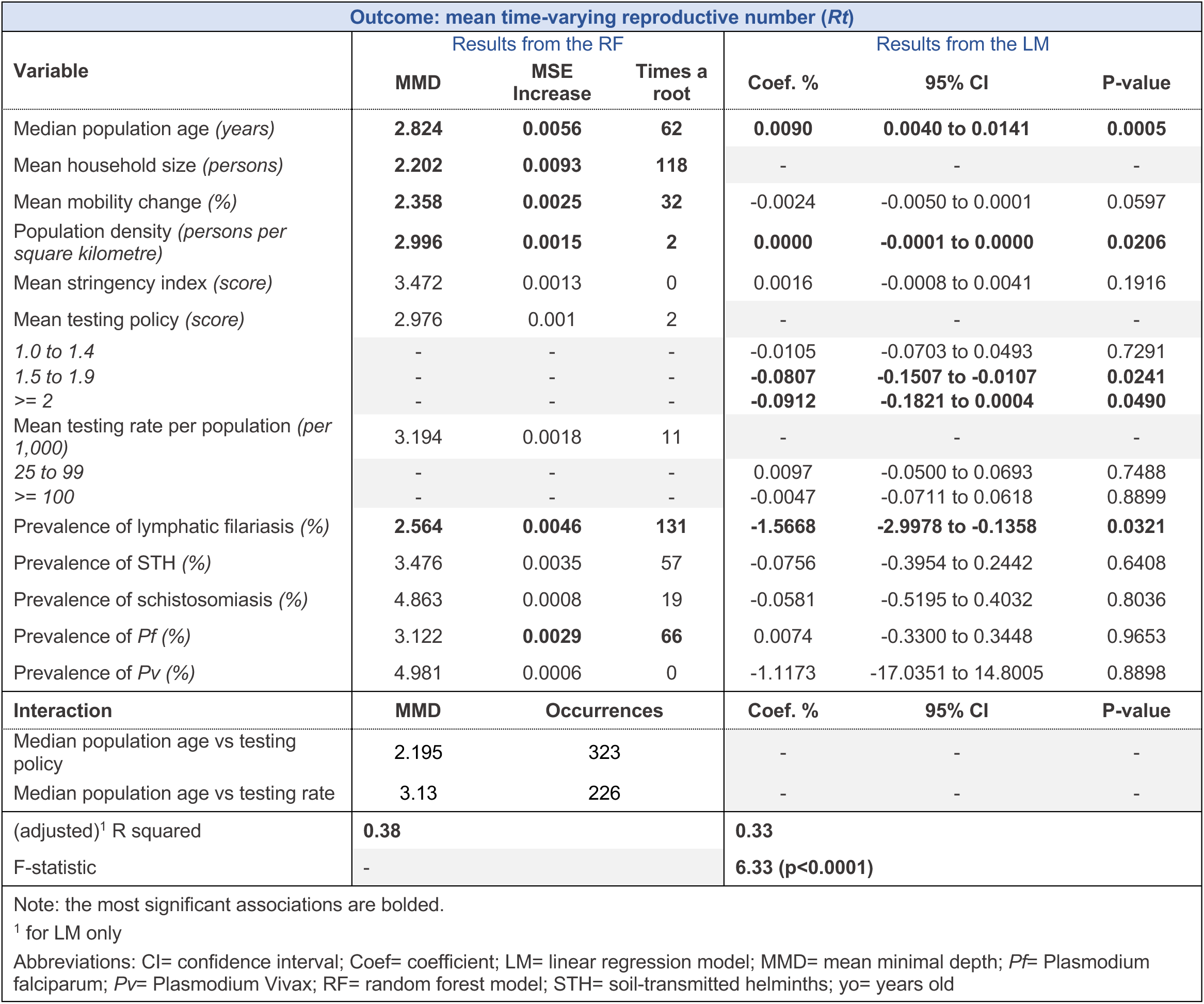
Results from random forest regression (RF) and multivariate linear regression (LM) models with all potential exposures for mean reproductive number (*Rt*)

| Outcome: mean time-varying reproductive number ($R_t$) | | | | | | |
| --- | --- | --- | --- | --- | --- | --- |
| Variable | Results from the RF |  |  | Results from the LM |  |  |
|  | MMD | MSE Increase | Times a root | Coef. % | 95% CI | P-value |
| Median population age (*years*) | **2.824** | **0.0056** | **62** | **0.0090** | **0.0040 to 0.0141** | **0.0005** |
| Mean household size (*persons*) | **2.202** | **0.0093** | **118** | - | - | - |
| Mean mobility change (%) | **2.358** | **0.0025** | **32** | -0.0024 | -0.0050 to 0.0001 | 0.0597 |
| Population density (*persons per square kilometre*) | **2.996** | **0.0015** | **2** | **0.0000** | **-0.0001 to 0.0000** | **0.0206** |
| Mean stringency index (*score*) | 3.472 | 0.0013 | 0 | 0.0016 | -0.0008 to 0.0041 | 0.1916 |
| Mean testing policy (*score*) | 2.976 | 0.001 | 2 | - | - | - |
| 1.0 to 1.4 | - | - | - | -0.0105 | -0.0703 to 0.0493 | 0.7291 |
| 1.5 to 1.9 | - | - | - | **-0.0807** | **-0.1507 to -0.0107** | **0.0241** |
| >= 2 | - | - | - | **-0.0912** | **-0.1821 to 0.0004** | **0.0490** |
| Mean testing rate per population (*per 1,000*) | 3.194 | 0.0018 | 11 | - | - | - |
| 25 to 99 | - | - | - | 0.0097 | -0.0500 to 0.0693 | 0.7488 |
| >= 100 | - | - | - | -0.0047 | -0.0711 to 0.0618 | 0.8899 |
| Prevalence of lymphatic filariasis (%) | **2.564** | **0.0046** | **131** | **-1.5668** | **-2.9978 to -0.1358** | **0.0321** |
| Prevalence of STH (%) | 3.476 | 0.0035 | 57 | -0.0756 | -0.3954 to 0.2442 | 0.6408 |
| Prevalence of schistosomiasis (%) | 4.863 | 0.0008 | 19 | -0.0581 | -0.5195 to 0.4032 | 0.8036 |
| Prevalence of *Pf* (%) | 3.122 | **0.0029** | **66** | 0.0074 | -0.3300 to 0.3448 | 0.9653 |
| Prevalence of *Pv* (%) | 4.981 | 0.0006 | 0 | -1.1173 | -17.0351 to 14.8005 | 0.8898 |
| Interaction | MMD | Occurrences |  | Coef. % | 95% CI | P-value |
| Median population age vs testing policy | 2.195 | 323 |  | - | - | - |
| Median population age vs testing rate | 3.13 | 226 |  | - | - | - |
| (adjusted)<sup>1</sup> R squared | **0.38** |  |  | **0.33** |  |  |
| F-statistic | - |  |  | **6.33 (p<0.0001)** |  |  |

Note: the most significant associations are bolded.
<sup>1</sup> for LM only
Abbreviations: CI= confidence interval; Coef= coefficient; LM= linear regression model; MMD= mean minimal depth; *Pf*= Plasmodium falciparum; *Pv*= Plasmodium Vivax; RF= random forest model; STH= soil-transmitted helminths; yo= years old

**Table S5.** Results from random forest regression (RF) and multivariate linear regression (LM) models with all potential exposures for age-standardised median age of observed cases

| Outcome: age-standardised median age of observed cases |  |  |  |  |  |  |
| --- | --- | --- | --- | --- | --- | --- |
| Variable | Results from the RF |  |  | Results from the LM |  |  |
|  | MMD | MSE Increase | Times a root | Coef. % | 95% CI | P-value |
| Median population age (*years*) | **2.144** | **6.2533** | **106** | **-0.6676** | **-1.2665 to -0.0688** | **0.0300** |
| Mean mobility change (%) | **2.537** | **1.4986** | **28** | - | - | - |
| -29 to -20 | - | - | - | 0.6044 | -3.9411 to 5.1500 | 0.7886 |
| -19 to -10 | - | - | - | 0.8265 | -3.4628 to 5.1157 | 0.6978 |
| -10 or less | - | - | - | **6.1251** | **0.5028 to 11.7475** | **0.0336** |
| Mean stringency index (*score*) | 2.933 | 0.8334 | 7 | 0.0190 | -0.2348 to 0.2728 | 0.8799 |
| Mean testing policy (*score*) | 3.107 | 0.7947 | 29 | - | - | - |
| 1.0 to 1.4 | - | - | - | 1.8291 | -2.4798 to 6.1379 | 0.3944 |
| 1.5 to 1.9 | - | - | - | 1.7326 | -2.8923 to 6.3575 | 0.4517 |
| >= 2 | - | - | - | 1.8407 | -4.2195 to 7.8910 | 0.5405 |
| Mean testing rate per population (*per 1,000*) | **2.434** | **2.0958** | **76** | - | - | - |
| 25 to 99 | - | - | - | 1.6929 | -3.1806 to 6.5664 | 0.4850 |
| >= 100 | - | - | - | -2.6100 | -8.0421 to 2.8222 | 0.3357 |
| Proportion of population at increased risk (%) | **2.169** | **4.0191** | **85** | -0.0839 | -0.5728 to 0.4050 | 0.7294 |
| Prevalence of lymphatic filariasis (%) | 5.352 | 0.3424 | 18 | - | - | - |
| 0.1 to 2.4 | - | - | - | -2.3861 | -7.8797 to 3.0976 | 0.3828 |
| >= 2.5 | - | - | - | 0.5672 | -7.3758 to 8.5102 | 0.8855 |
| Prevalence of STH (%) | 3.179 | **2.2431** | **80** | - | - | - |
| 0.1 to 9.9 | - | - | - | -6.8029 | -13.8022 to 0.1964 | 0.0564 |
| 10.0 to 19.9 | - | - | - | -3.7679 | -12.2819 to 4.7461 | 0.3748 |
| >= 20.0 | - | - | - | -6.6602 | -16.9021 to 3.5816 | 0.1951 |
| Prevalence of schistosomiasis (%) | 4.926 | 0.9941 | 37 | - | - | - |
| 0.1% to 4.9% | - | - | - | 3.7039 | -2.0660 to 9.4737 | 0.2008 |
| >= 5.0% | - | - | - | 1.6109 | -5.6544 to 8.8762 | 0.6551 |
| Prevalence of *Pf* (%) | 3.573 | 0.1091 | 32 | - | - | - |
| 0.1% to 4.9% | - | - | - | -3.2222 | -8.8398 to 2.3955 | 0.2519 |
| >= 5.0% | - | - | - | -2.1715 | -12.2269 to 7.8838 | 0.6635 |
| Prevalence of *Pv* (%) | 4.894 | -0.0549 | 2 | - | - | - |
| > 0% | - | - | - | 2.4024 | -2.1081 to 6.9128 | 0.2867 |
| Interaction | MMD | Occurrences |  | Coef. % | 95% CI | P-value |
| Median population age vs mean mobility change | 2.210 | 206 |  | - | - | - |
| Median population age vs mean stringency index | 1.980 | 201 |  | - | - | - |
| Median population age vs proportion at increased risk | 1.376 | 251 |  | - | - | - |
| (adjusted)<sup>1</sup> R squared | **0.25** |  |  | **0.37** |  |  |
| F-statistic | - |  |  | **2.53 (p < 0.01)** |  |  |

Note: the most significant associations are bolded.
<sup>1</sup> for LM only
Abbreviations: CI= confidence interval; Coef= coefficient; LM= linear regression model; MMD= mean minimal depth; *Pf*= Plasmodium falciparum; *Pv*= Plasmodium Vivax; RF= random forest model; STH= soil-transmitted helminths; yo= years old

**Table S6.** Results from random forest regression (RF) and multivariate linear regression (LM) models with all potential exposures for age-standardised median age of observed deaths

| Outcome: age-standardised median age of observed deaths |  |  |  |  |  |  |
| --- | --- | --- | --- | --- | --- | --- |
| Variable | Results from the RF |  |  | Results from the LM |  |  |
|  | MMD | MSE Increase | Times a root | Coef. % | 95% CI | P-value |
| Median population age (*years*) | **1.768** | **9.8945** | **119** | -0.1302 | -1.1497 to 0.8893 | 0.7908 |
| Mean mobility change (%) | 2.891 | -0.0752 | 8 | - | - | - |
| -29 to -20 | - | - | - | -0.3183 | -6.3763 to 5.7388 | 0.9130 |
| -19 to -10 | - | - | - | -1.0371 | -6.9453 to 4.8710 | 0.7157 |
| -10 or less | - | - | - | -1.8881 | -9.0188 to 5.2426 | 0.5837 |
| Mean stringency index (*score*) | **1.848** | **8.9399** | **92** | -0.2488 | -0.5791 to 0.0816 | 0.1305 |
| Mean testing policy (*score*) | 2.677 | -0.0939 | 11 | - | - | - |
| 1.0 to 1.4 | - | - | - | 0.6690 | -5.8649 to 7.2028 | 0.8315 |
| 1.5 to 1.9 | - | - | - | 0.7056 | -6.4011 to 7.8123 | 0.8366 |
| >= 2 | - | - | - | 3.5438 | -4.8425 to 11.9301 | 0.3851 |
| Mean testing rate per population (*per 1,000*) | **1.946** | **6.0048** | **71** | - | - | - |
| 25 to 99 | - | - | - | -4.1627 | -10.6720 to 2.3466 | 0.3851 |
| >= 100 | - | - | - | 0.8141 | -6.3255 to 7.9536 | 0.1950 |
| Proportion of population at increased risk (%) | **1.727** | **6.5484** | **68** | -0.6593 | -1.6830 to 0.3644 | -0.1920 |
| Prevalence of lymphatic filariasis (%) | 4.724 | 0.0554 | 6 | - | - | - |
| 0.1% to 2.4% | - | - | - | -10.1806 | -26.1029 to 5.7417 | 0.1950 |
| >= 2.5% | - | - | - | -9.6474 | -31.9355 to 12.6407 | 0.3739 |
| Prevalence of STH (%) | 3.208 | **5.5241** | **87** | - | - | - |
| 0.1% to 9.9% | - | - | - | -0.3850 | -8.5390 to 7.7690 | 0.9218 |
| 10.0% to 19.9% | - | - | - | 1.1088 | -11.8046 to 14.0223 | 0.8584 |
| >= 20.0% | - | - | - | 2.5237 | -12.4134 to 17.4607 | 0.7259 |
| Prevalence of schistosomiasis (%) | 4.320 | 0.6929 | 18 | - | - | - |
| 0.1% to 4.9% | - | - | - | 9.0544 | -8.3629 to 26.4717 | 0.2880 |
| >= 5.0% | - | - | - | 10.0198 | -5.1538 to 25.1934 | 0.1815 |
| Prevalence of *Pf* (%) | 3.666 | 0.9885 | 18 | - | - | - |
| 0.1% to 4.9% | - | - | - | -8.8861 | -21.9717 to 4.1994 | 0.1701 |
| >= 5.0% | - | - | - | -8.0450 | -25.5108 to 9.4209 | 0.3448 |
| Prevalence of *Pv* (%) | 4.269 | 0.0061 | 2 | - | - | - |
| > 0% | - | - | - | 1.5756 | -5.6915 to 8.8428 | 0.6531 |
| Interaction | MMD | Occurrences |  | Coef. % | 95% CI | P-value |
| Median population age x mean mobility change | 2.005 | 105 |  | - | - | - |
| Median population age x mean stringency index | 1.496 | 171 |  | - | - | - |
| Median population age x proportion at increased risk | 1.923 | 162 |  | - | - | - |
| (adjusted)<sup>1</sup> R squared | **0.63** |  |  | **0.67** |  |  |
| F-statistic | - |  |  | **4.61 (p< 0.01)** |  |  |

Note: the most significant associations are bolded.
<sup>1</sup> for LM onlyAbbreviations: CI= confidence interval; Coef= coefficient; LM= linear regression model; MMD= mean minimal depth; *Pf*= Plasmodium falciparum; *Pv*= Plasmodium Vivax; RF= random forest model; STH= soil-transmitted helminths; yo= years old

**Table S7.** Results from random forest regression (RF) and multivariate linear regression (LM) models with all potential exposures for crude case fatality ratio (CFR)

| Outcome: crude CFR |  |  |  |  |  |  |
| --- | --- | --- | --- | --- | --- | --- |
| Variable | Results from the RF |  |  | Results from the LM |  |  |
|  | MMD | MSE Increase | Times a root | Coef. % | 95% CI | P-value |
| Median population age (years) | 2.518 | 0.0002 | 50 | 0.0301 | -0.0189 to 0.0792 | 0.2268 |
| Mean mobility change (%) | - | - | - | - | - | - |
| -29 to -20 | - | - | - | -0.0760 | -0.4931 to 0.3412 | 0.7194 |
| -19 to -10 | - | - | - | -0.2997 | -0.7580 to 0.1585 | 0.1981 |
| -10 or less | - | - | - | 0.0485 | -0.6525 to 0.5554 | 0.8740 |
| Mean stringency index (score) | 2.904 | 0.0000 | 65 | 0.0093 | -0.0084 to 0.0270 | 0.2995 |
| Mean testing policy (score) | 1.998 | 0.0002 | 119 | - | - | - |
| 1.0 to 1.4 | - | - | - | -0.2809 | -0.6835 to 0.1216 | 0.1698 |
| 1.5 to 1.9 | - | - | - | -0.7011 | -1.1693 to -0.2330 | 0.0036 |
| >= 2 | - | - | - | -0.6057 | -1.2208 to 0.0095 | 0.0536 |
| Mean testing rate per population (per 1,000) | 3.340 | 0.0001 | 40 | - | - | - |
| 25 to 99 | - | - | - | -0.2566 | -0.6673 to 0.1540 | 0.2187 |
| >= 100 | - | - | - | -0.3369 | -0.8422 to 0.1684 | 0.1896 |
| Proportion of population at increased risk (%) | 2.100 | 0.0001 | 78 | -0.0345 | -0.0905 to 0.0214 | 0.2240 |
| Prevalence of lymphatic filariasis (%) | 4.260 | 0.0001 | 50 | - | - | - |
| 0.1 to 2.4 | - | - | - | 0.2595 | -0.2516 to 0.7706 | 0.3172 |
| >= 2.5 | - | - | - | 0.5025 | -0.1300 to 1.1349 | 0.1185 |
| Prevalence of STH (%) | 3.476 | 0.0001 | 35 | - | - | - |
| 0.1 to 9.9 | - | - | - | -0.5263 | -1.1182 to 0.0656 | 0.0810 |
| 10.0 to 19.9 | - | - | - | -0.4017 | -1.1499 to 0.3465 | 0.2903 |
| >= 20.0 | - | - | - | -0.9144 | -1.7933 to -0.0356 | 0.0415 |
| Prevalence of schistosomiasis (%) | 5.241 | 0.0002 | 14 | - | - | - |
| 0.1 to 4.9 | - | - | - | 0.1885 | -0.2651 to 0.6420 | 0.4128 |
| >= 5.0 | - | - | - | 0.1840 | -0.3700 to 0.7379 | 0.5125 |
| Prevalence of Pf (%) | 4.443 | 0.0002 | 18 | - | - | - |
| 0.1 to 4.9 | - | - | - | 0.0952 | -0.4515 to 0.6419 | 0.7311 |
| >= 5.0 | - | - | - | 0.0484 | -0.7504 to 0.8473 | 0.9048 |
| Prevalence of Pv (%) | 4.604 | 0.0001 | 31 | - | - | - |
| > 0 | - | - | - | 0.3058 | -0.1761 to 0.7876 | 0.2117 |
| **Interaction** | **MMD** | **Occurrences** |  | **Coef. %** | **95% CI** | **P-value** |
| Median population age x mean mobility change | n/a | 0 |  | - | - | - |
| Median population age x mean stringency index | 1.920 | 326 |  | - | - | - |
| (adjusted)<sup>1</sup> R squared | -0.06 |  |  | 0.07 |  |  |
| F-statistic | - |  |  | 1.54 (p = 0.0725) |  |  |
| <sup>1</sup> for LM only |  |  |  |  |  |  |
| Abbreviations: CFR= case-fatality ratio; CI= confidence interval; Coef= coefficient; LM= linear regression model; MMD= mean minimal depth; Pf= Plasmodium falciparum; Pv= Plasmodium Vivax; RF= random forest model; STH= soil-transmitted helminths; yo= years old |  |  |  |  |  |  |

**Table S8.** Results from random forest regression (with imputation) and multivariate linear regression models with all potential exposures for age-standardised case fatality ratio (CFR)

| Outcome: age-standardised CFR |  |  |  |  |  |  |
| --- | --- | --- | --- | --- | --- | --- |
| Variable | Results from the RF |  |  | Results from the LM |  |  |
|  | MMD | MSE Increase | Times a root | Coef. % | 95% CI | P-value |
| Median population age (*years*) | **1.851** | 0.0000 | **105** | 0.1169 | -0.3255 to 0.5593 | 0.5649 |
| Mean mobility change (%) | - | - | - | - | - | - |
| -29 to -20 | - | - | - | 2.0643 | -0.9257 to 5.0543 | 0.1528 |
| -19 to -10 | - | - | - | 1.5586 | -0.9079 to 4.0252 | 0.1866 |
| -10 or less | - | - | - | 1.9032 | -0.5630 to 4.3693 | 0.1148 |
| Mean stringency index (*score*) | 2.950 | 0.0000 | 28 | 0.0471 | -0.1027 to 0.1970 | 0.4948 |
| Mean testing policy (*score*) | 2.675 | 0.0000 | 39 | - | - | - |
| 1.0 to 1.4 | - | - | - | 0.2622 | -1.5290 to 2.0534 | 0.7482 |
| 1.5 to 1.9 | - | - | - | -0.0100 | -2.5425 to 2.5225 | 0.9931 |
| >= 2 | - | - | - | 0.1802 | -2.6877 to 3.0482 | 0.8901 |
| Mean testing rate per population (*per 1,000*) | 3.221 | 0.0000 | 23 | - | - | - |
| 25 to 99 | - | - | - | 0.6494 | -1.6477 to 2.9466 | 0.5384 |
| >= 100 | - | - | - | 0.9713 | -1.2607 to 3.2033 | 0.3506 |
| Proportion of population at increased risk (%) | **2.403** | 0.0000 | **68** | 0.1247 | -0.1595 to 0.4089 | 0.3468 |
| Prevalence of lymphatic filariasis (%) | 4.436 | 0.0000 | 29 | - | - | - |
| 0.1 to 2.4 | - | - | - | 2.1934 | -2.6004 to 6.9871 | 0.3277 |
| >= 2.5 | - | - | - | 5.3592 | -2.8973 to 13.6157 | 0.1761 |
| Prevalence of STH (%) | 3.634 | 0.0000 | 42 | - | - | - |
| 0.1 to 9.9 | - | - | - | -0.7032 | -3.2564 to 1.8599 | 0.5487 |
| 10.0 to 19.9 | - | - | - | -1.7365 | -6.3547 to 2.8817 | 0.4170 |
| >= 20.0 | - | - | - | -2.0912 | -7.2535 to 3.0711 | 0.3834 |
| Prevalence of schistosomiasis (%) | 4.675 | 0.0000 | 52 | - | - | - |
| 0.1 to 4.9 | - | - | - | -2.3752 | -7.5956 to 2.8453 | 0.3302 |
| >= 5.0 | - | - | - | -3.1224 | -8.7548 to 2.5101 | 0.2414 |
| Prevalence of *Pf* (%) | 2.888 | 0.0000 | 91 | - | - | - |
| 0.1 to 4.9 | - | - | - | 4.4215 | -2.9675 to 11.8105 | 0.2089 |
| >= 5.0 | - | - | - | 3.9715 | -4.7797 to 12.7228 | 0.3314 |
| Prevalence of *Pv* (%) | 3.050 | 0.0000 | 37 | - | - | - |
| > 0 | - | - | - | 0.9920 | -1.2756 to 3.2596 | 0.3482 |
| Interaction | MMD | Occurrences |  | Coef. % | 95% CI | P-value |
| Median population age x mean mobility change | n/a | 0 |  | - | - | - |
| Median population age x mean stringency index | 0.930 | 128 |  | - | - | - |
| (adjusted)<sup>1</sup> R squared | **-0.21** |  |  | 0.01 |  |  |
| F-statistic | - |  |  | 1.02 (p = 0.5161) |  |  |
| <sup>1</sup> for LM only<br>Abbreviations: CFR= case-fatality ratio; CI= confidence interval; Coef= coefficient; LM= linear regression model; MMD= mean minimal depth; *Pf*= Plasmodium falciparum; *Pv*= Plasmodium Vivax; RF= random forest model; STH= soil-transmitted helminths; yo= years old |  |  |  |  |  |  |

**Table S9.** Results from random forest regression (with imputation) and multivariate linear regression models with all potential exposures for incidence-standardised case fatality ratio (CFR)

| Outcome: incidence-standardised CFR |  |  |  |  |  |  |
| --- | --- | --- | --- | --- | --- | --- |
| Variable | Results from the RF |  |  | Results from the LM |  |  |
|  | MMD | MSE Increase | Times a root | Coef. % | 95% CI | P-value |
| Median population age (*years*) | 1.884 | 0.0000 | 103 | 0.0042 | -0.0012 to 0.0095 | 0.1086 |
| Mean mobility change (%) | - | - | - | - | - | - |
| -29 to -20 | - | - | - | 0.0537 | -0.0149 to 0.0925 | 0.0128 |
| -19 to -10 | - | - | - | 0.0476 | -0.0147 to 0.0806 | 0.0103 |
| -10 or less | - | - | - | 0.0532 | -0.0165 to 0.0900 | 0.0102 |
| Mean stringency index (*score*) | 2.771 | 0.0000 | 24 | 0.0014 | 0.0003 to 0.0032 | 0.0987 |
| Mean testing policy (*score*) | 2.750 | 0.0000 | 46 | - | - | - |
| 1.0 to 1.4 | - | - | - | -0.0086 | -0.0323 to 0.0152 | 0.4306 |
| 1.5 to 1.9 | - | - | - | -0.0250 | -0.0609 to 0.0109 | 0.1474 |
| >= 2 | - | - | - | -0.0189 | -0.0546 to 0.0168 | 0.2563 |
| Mean testing rate per population (*per 1,000*) | 3.279 | 0.0000 | 12 | - | - | - |
| 25 to 99 | - | - | - | 0.0055 | -0.0216 to 0.0327 | 0.6503 |
| >= 100 | - | - | - | 0.0300 | 0.0031 to 0.0570 | 0.4898 |
| Proportion of population at increased risk (%) | 2.360 | 0.0000 | 65 | 0.0034 | -0.0005 to 0.0072 | 0.0774 |
| Prevalence of lymphatic filariasis (%) | 4.282 | 0.0000 | 24 | - | - | - |
| 0.1 to 2.4 | - | - | - | 0.0290 | -0.0263 to 0.0842 | 0.2611 |
| >= 2.5 | - | - | - | 0.0369 | -0.0698 to 0.1435 | 0.4484 |
| Prevalence of STH (%) | 3.153 | 0.0000 | 94 | - | - | - |
| 0.1 to 9.9 | - | - | - | -0.0058 | -0.0354 to 0.0238 | 0.6622 |
| 10.0 to 19.9 | - | - | - | -0.0192 | -0.0724 to 0.0340 | 0.4291 |
| >= 20.0 | - | - | - | 0.0116 | -0.0699 to 0.0931 | 0.7510 |
| Prevalence of schistosomiasis (%) | 4.380 | 0.0000 | 26 | - | - | - |
| 0.1 to 4.9 | - | - | - | -0.0745 | -0.1552 to 0.0063 | 0.0662 |
| >= 5.0 | - | - | - | -0.0620 | -0.1279 to 0.0040 | 0.0623 |
| Prevalence of *Pf* (%) | 3.086 | 0.0000 | 73 | - | - | - |
| 0.1 to 4.9 | - | - | - | 0.1165 | 0.0231 to 0.2100 | 0.0206 |
| >= 5.0 | - | - | - | 0.1507 | 0.0338 to 0.2676 | 0.0178 |
| Prevalence of *Pv* (%) | 3.719 | 0.0000 | 33 | - | - | - |
| > 0 | - | - | - | 0.0167 | -0.097 to 0.0430 | 0.1824 |
| Interaction | MMD | Occurrences |  | Coef. % | 95% CI | P-value |
| Median population age x mean mobility change | n/a | 0 |  | - | - | - |
| Median population age x mean stringency index | 1.351 | 127 |  | - | - | - |
| (adjusted)<sup>1</sup> R squared | -0.02 |  |  | 0.61 |  |  |
| F-statistic | - |  |  | 3.18 (p = 0.0481) |  |  |
| <sup>1</sup> for LM only |  |  |  |  |  |  |
| Abbreviations: CFR= case-fatality ratio; CI= confidence interval; Coef= coefficient; LM= linear regression model; MMD= mean minimal depth; *Pf*= Plasmodium falciparum; *Pv*= Plasmodium Vivax; RF= random forest model; STH= soil-transmitted helminths; yo= years old |  |  |  |  |  |  |

**Table S10.** Result from random forest regression (RF) and multivariate linear regression (LM) models with all potential exposures for mean reproductive number (Rt) for the WHO AFRO region

| Outcome: mean time-varying reproductive number (*R*t) – AFRO region |  |  |  |  |  |  |
| --- | --- | --- | --- | --- | --- | --- |
| Variable | Results from the RF |  |  | Results from the LM |  |  |
|  | MMD | MSE Increase | Times a root | Coef. % | 95% CI | P-value |
| Median population age (*years*) | 3.545 | -0.0001 | 21 | 0.0101 | -0.0251 to 0.0452 | 0.5548 |
| Mean household size (*persons*) | **2.136** | **0.0030** | **92** | - | - | - |
| Mean mobility change (%) | **3.222** | **0.0010** | **60** | 0.0033 | -0.0038 to 0.0105 | 0.3371 |
| Population density (*persons per square kilometre*) | 3.691 | -0.0003 | 22 | 0.0003 | -0.0004 to 0.0011 | 0.3561 |
| Mean stringency index (*score*) | 3.425 | 0.0003 | 21 | 0.0023 | -0.0035 to 0.0081 | 0.4116 |
| Mean testing policy (*score*) | 2.986 | -0.0003 | 25 | - | - | - |
| 1.0 to 1.4 | - | - | - | -0.1167 | -0.2479 to 0.0145 | 0.0781 |
| 1.5 to 1.9 | - | - | - | -0.0648 | -0.2692 to 0.1396 | 0.5136 |
| >= 2 | - | - | - | -0.1512 | -0.4378 to 0.1353 | 0.2822 |
| Mean testing rate per population (*per 1,000*) | 3.581 | 0.0000 | 12 | - | - | - |
| 25 to 99 | - | - | - | 0.0584 | -0.0858 to 0.2025 | 0.4062 |
| >= 100 | - | - | - | -0.0978 | -0.4720 to 0.2763 | 0.5896 |
| Prevalence of lymphatic filariasis (%) | **1.766** | **0.0046** | **144** | **-2.6287** | **-4.7969 to -0.4605** | **0.0202** |
| Prevalence of STH (%) | 4.429 | 0.0000 | 8 | 0.5238 | -0.3942 to 1.4417 | 0.2462 |
| Prevalence of schistosomiasis (%) | 4.132 | -0.0002 | 19 | -0.4160 | -1.1614 to 0.3294 | 0.2562 |
| Prevalence of *Pf* (%) | **3.008** | **0.0011** | **76** | **-0.4841** | **-0.9459 to -0.0222** | **0.0410** |
| Prevalence of *Pv* (%) | 6.208 | 0.0000 | 0 | -34.4310 | -96.0170 to 27.1550 | 0.2555 |
| Interaction | MMD | Occurrences |  | Coef. % | 95% CI | P-value |
| Median population age vs testing policy | 2.0206 | 97 |  | - | - | - |
| Median population age vs testing rate | 2.0494 | 92 |  | - | - | - |
| (adjusted)<sup>1</sup> R squared | **0.19** |  |  | 0.19 |  |  |
| F-statistic | - |  |  | 1.53 (p= 0.1953) |  |  |
| Note: the most significant associations are bolded. |  |  |  |  |  |  |
| <sup>1</sup> for LM only |  |  |  |  |  |  |
| Abbreviations: CI= confidence interval; Coef= coefficient; LM= linear regression model; MMD= mean minimal depth; *Pf*= Plasmodium falciparum; *Pv*= Plasmodium Vivax; RF= random forest model; STH= soil-transmitted helminths; yo= years old |  |  |  |  |  |  |

**Table S11.** Result from reduced multivariate linear regression (LM) model for mean reproductive number (Rt) for the WHO AFRO region

| Outcome: mean time-varying reproductive number (*R*t) – AFRO region |  |  |  |
| --- | --- | --- | --- |
| Variable | Results from the reduced LM |  |  |
|  | Coef. % | 95% CI | P-value |
| Prevalence of lymphatic filariasis (%) | -2.6132 | -4.0060 to -1.2205 | 0.0006 |
| Population density (*persons per square kilometre*) | 0.0003 | -0.0001 to 0.0006 | 0.1988 |
| Mean stringency index (*score*) | -0.0003 | -0.0043 to 0.0038 | 0.8939 |
| (adjusted) R squared | 0.31 |  |  |
| F-statistic | 6.38 (p= 0.0016) |  |  |
| Abbreviations: CI= confidence interval; Coef= coefficient; LM= linear regression model; |  |  |  |

**Table S12.** Result from random forest regression (RF) and multivariate linear regression (LM) models with all potential exposures for crude case fatality ratio (CFR) for the WHO AFRO region

| Outcome: crude CFR |  |  |  |  |  |  |
| --- | --- | --- | --- | --- | --- | --- |
| Variable | Results from the RF |  |  | Results from the LM |  |  |
|  | MMD | MSE Increase | Times a root | Coef. % | 95% CI | P-value |
| Median population age (*years*) | 2.086 | 0.0000 | 98 | 0.056 | -0.159 to 0.272 | 0.5834 |
| Mean mobility change (%) | - | - | - | - | - | - |
| -29 to -20 | - | - | - | 0.345 | -0.733 to 1.423 | 0.5039 |
| -19 to -10 | - | - | - | -0.340 | -1.582 to 0.902 | 0.5666 |
| -10 or less | - | - | - | -0.117 | -1.600 to 1.366 | 0.8680 |
| Mean stringency index (*score*) | 3.642 | 0.0000 | 38 | -0.002 | -0.057 to 0.052 | 0.9331 |
| Mean testing policy (*score*) | 2.936 | 0.0000 | 39 | - | - | - |
| 1.0 to 1.4 | - | - | - | -0.820 | -1.933 to 0.294 | 0.1366 |
| 1.5 to 1.9 | - | - | - | -0.445 | -1.880 to 0.991 | 0.5171 |
| >= 2 | - | - | - | -2.193 | -3.918 to -0.469 | 0.0163 |
| Mean testing rate per population (*per 1,000*) | 3.941 | 0.0000 | 41 | - | - | - |
| 25 to 99 | - | - | - | 0.586 | -0.427 to 1.599 | 0.2353 |
| Proportion of population at increased risk (%) | 4.035 | 0.0000 | 23 | -0.078 | -0.207 to 0.050 | 0.2114 |
| Prevalence of lymphatic filariasis (%) | 3.410 | 0.0000 | 87 | - | - | - |
| 0.1 to 2.4 | - | - | - | 1.024 | -0.799 to 2.847 | 0.2484 |
| >= 2.5 | - | - | - | 1.006 | -0.863 to 2.876 | 0.2675 |
| Prevalence of STH (%) | 2.612 | 0.0000 | 103 | - | - | - |
| 10.0 to 19.9 | - | - | - | 1.233 | -0.389 to 2.855 | 0.1252 |
| >= 20.0 | - | - | - | 0.988 | -0.560 to 2.535 | 0.1926 |
| Prevalence of schistosomiasis (%) | 3.428 | 0.0000 | 39 | - | - | - |
| 0.1 to 4.9 | - | - | - | -0.510 | -2.037 to 1.018 | 0.4860 |
| >= 5.0 | - | - | - | -0.808 | -2.476 to 0.859 | 0.3162 |
| Prevalence of *Pf* (%) | 3.963 | 0.0000 | 27 | - | - | - |
| 0.1 to 4.9 | - | - | - | -0.415 | -2.499 to 1.669 | 0.6758 |
| >= 5.0 | - | - | - | -0.795 | -3.729 to 2.139 | 0.5704 |
| Prevalence of *Pv* (%) | 6.889 | 0.0000 | 5 | - | - | - |
| > 0 | - | - | - | -1.394 | -3.460 to -0.672 | 0.1700 |
| Interaction | MMD | Occurrences |  | Coef. % | 95% CI | P-value |
| Median population age x mean mobility change | n/a | 0 |  | - | - | - |
| Median population age x mean stringency index | 1.605 | 162 |  | - | - | - |
| (adjusted)<sup>1</sup> R squared | -0.03 |  |  | 0.05 |  |  |
| F-statistic | - |  |  | 1.10(p = 0.4359) |  |  |
| <sup>1</sup> for LM only<br>Abbreviations: CFR= case-fatality ratio; CI= confidence interval; Coef= coefficient; LM= linear regression model; MMD= mean minimal depth; *Pf*= Plasmodium falciparum; *Pv*= Plasmodium Vivax; RF= random forest model; STH= soil-transmitted helminths; yo= years old |  |  |  |  |  |  |

**Table S13.**
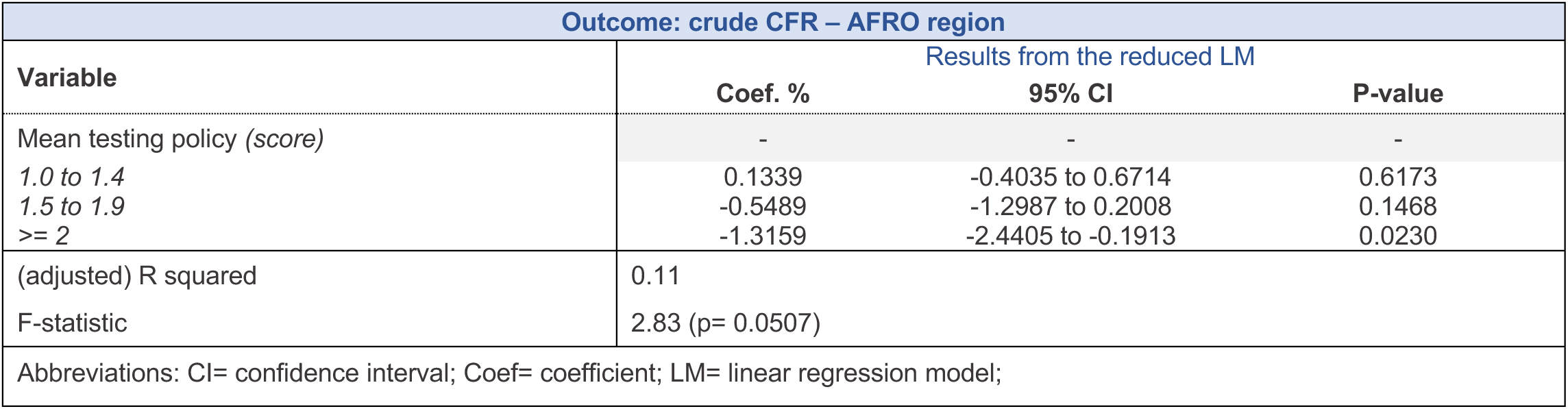
Result from reduced multivariate linear regression (LM) model for crude CFR for the WHO AFRO region

| Outcome: crude CFR – AFRO region |  |  |  |
| --- | --- | --- | --- |
| Variable | Results from the reduced LM |  |  |
|  | Coef. % | 95% CI | P-value |
| Mean testing policy (*score*) | - | - | - |
| 1.0 to 1.4 | 0.1339 | -0.4035 to 0.6714 | 0.6173 |
| 1.5 to 1.9 | -0.5489 | -1.2987 to 0.2008 | 0.1468 |
| >= 2 | -1.3159 | -2.4405 to -0.1913 | 0.0230 |
| (adjusted) R squared | 0.11 |  |  |
| F-statistic | 2.83 (p= 0.0507) |  |  |
| Abbreviations: CI= confidence interval; Coef= coefficient; LM= linear regression model; |  |  |  |

